# What does the COVID-19 pandemic mean for the next decade of onchocerciasis control and elimination?

**DOI:** 10.1101/2020.10.26.20219733

**Authors:** Jonathan I.D. Hamley, David J. Blok, Martin Walker, Philip Milton, Adrian D. Hopkins, Louise C. Hamill, Philip Downs, Sake J. de Vlas, Wilma A. Stolk, Maria-Gloria Basáñez

## Abstract

**Background:** Mass drug administration (MDA) of ivermectin for onchocerciasis has been disrupted by the SARS-CoV-2 (COVID-19) pandemic. Mathematical modelling can help predict how missed/delayed MDA will affect short-term epidemiological trends and elimination prospects by 2030.

**Methods:** Two onchocerciasis transmission models (EPIONCHO-IBM and ONCHOSIM) are used to simulate microfilarial prevalence trends, elimination probabilities, and age-profiles of *Onchocerca volvulus* microfilarial prevalence and intensity, for different treatment histories and transmission settings, assuming no interruption, a 1-year (2020) or 2-year (2020–2021) interruption. Biannual MDA or increased coverage upon MDA resumption are investigated as remedial strategies.

**Results:** Programmes with shorter MDA histories and settings with high pre-intervention endemicity will be the most affected. Biannual MDA is more effective than increasing coverage for mitigating COVID-19’s impact on MDA. Programmes which had already switched to biannual MDA should be minimally affected. In high transmission settings with short treatment history, a 2-year interruption could lead to increased microfilarial load in children (EPIONCHO-IBM) or adults (ONCHOSIM).

**Conclusions:** Programmes with shorter (annual MDA) treatment histories should be prioritised for remedial biannual MDA. Increases in microfilarial load could have short- and long-term morbidity and mortality repercussions. These results can guide decision-making to mitigate the impact of COVID-19 on onchocerciasis elimination.

## Introduction

The SARS-CoV-2 virus (COVID-19) pandemic has led to severe disruptions to routine public health services on a global scale. These disruptions are expected to be particularly pronounced in low- and middle-income countries due to already under-resourced healthcare systems. On April 1^st^ 2020, the World Health Organization (WHO) advised that mass drug administration (MDA) and epidemiological surveys for neglected tropical diseases (NTDs) tackled by preventive chemotherapy and transmission control (PCT) should be postponed.^1^ Updated guidance was released on July 27^th^, which included a decision-making framework, allowing countries to restart routine MDA given careful risk assessment.^2^ Onchocerciasis is one such PCT disease, centred on ivermectin (Mectizan^®^) MDA, delivered annually in the majority of endemic countries in Africa.

Although so-called ‘lockdowns’ and delayed MDA might be effective in temporally reducing the transmission of the SARS-CoV-2 virus, the implications for both short- and long-term onchocerciasis transmission are less clear. The impact of the Ebola outbreaks in West Africa (2013–2016) on morbidity due to other diseases (such as tuberculosis and HIV) can provide some insight into the implications of withdrawing routine public health services.^3,4^ However, comparability is limited due to the stark differences in the scale and manifestations of Ebola and COVID-19. There are concerns that delaying MDA might increase onchocerciasis morbidity in the short term, and in the long term, undermine progress made towards the 2030 elimination of transmission (EOT) goals proposed in the recently launched WHO 2021– 2030 Road Map for NTDs.^5^ It is, therefore, important to quantify where (in terms of transmission setting and treatment history) the impact of postponements of ivermectin MDA for onchocerciasis will be most pronounced and to identify the most effective mitigation strategies to help affected programmes to get back on track. This will allow better planning and prioritisation of ivermectin distribution/treatment upon safe MDA resumption.

Delayed ivermectin MDA, or reduced treatment coverage, could result not only from population-wide lockdowns to reduce COVID-19 transmission and the resulting redirection or disruption of health services, but also from shortages in drug availability (due to slower production and supply chains, or exceeded drug shelf-life by the time MDA recommences).^6,7^ Deadlines for drug orders (typically by August for delivery in the following year), as required by the Mectizan Donation Program (MDP, the body providing oversight for ivermectin donation to endemic countries), might also be problematic. In addition to these challenges, programmes will have to be adaptive in the face of unforeseen setbacks which may emerge as MDA recommences, remedial strategies (such as increased MDA coverage and/or frequency) are attempted, and as the COVID-19 pandemic progresses.

Mathematical models of onchocerciasis transmission provide a useful predictive tool for understanding the impacts of ivermectin MDA interruptions on the short-term (increases in transmission intensity) and long-term (elimination prospects), as well as the potential benefit of remedial mitigation strategies to help programmes get back on track. Although the resurgence of helminth transmission is typically slower than that of viral, bacterial, and protozoan infections (due to differences in life-history), short-term increases in infection prevalence and intensity may increase onchocerciasis-associated morbidity.^8^ Delays in treating children who would otherwise receive ivermectin when turning 5 could result in higher microfilarial loads experienced early in life, which may impact health outcomes in later years.^9^

In this paper, we use the EPIONCHO-IBM and ONCHOSIM transmission models to: i) quantify where (in terms of transmission setting and treatment history) the impact of interruptions to ivermectin MDA for onchocerciasis will be most pronounced, allowing better planning and prioritisation of ivermectin distribution/treatment upon resumption, and ii) investigate how mitigation strategies based on increased frequency (biannual MDA) of increased coverage can help affected programmes to get back on track. Both models contributed insights to inform the WHO 2021–2030 NTD Roadmap,^10^ and provided preliminary but less comprehensive results for a report to the WHO on the impact of COVID-19 on NTD programmes.^11^

## Materials and Methods

### Models

We used two individual-based stochastic transmission models, namely EPIONCHO-IBM^12^ and ONCHOSIM^13^ to address seven questions concerning the effect of MDA disruptions due to COVID-19: (i) what is the impact of delaying treatment for one or two years upon microfilarial prevalence trends and probability of elimination by 2030?; (ii) which pre-intervention endemicity levels are particularly vulnerable?; (iii) how much more vulnerable are programmes with shorter treatment histories than those with longer ones?; (iv) which age groups will be most affected?; (v) are remedial strategies based on increasing treatment frequency (to biannual MDA) or increasing treatment coverage during the early stages of MDA resumption useful for mitigating any setbacks?; (vi) how will programmes that had already switched to biannual MDA be impacted?; and (vii) what will be the effect on (West African) countries whose NTD programmes had already been disrupted by the Ebola outbreak in 2014?

In the context of a 1-year interruption or 1 year of missed MDA, the terms ‘interruption’ or ‘missed’ are used to describe a situation in which the MDA round planned to take place 12 months after a successful round pre-COVID19 is not delivered (regardless of the calendar month in which treatment is usually distributed according to setting). ‘Remedial’ MDA implies that once MDA can safely resume,^2^ additional MDA (either an extra round or increased coverage within one round) is delivered. Hence, ‘remedial’ biannual MDA can be interpreted as ‘delayed treatment’ if a round of annual MDA is not delivered when planned but given in addition to the planned round in the following (not necessarily calendar) year. Although, for simplicity, modelling results are presented as if MDA were delivered at the beginning of each year, what is important is the duration between two consecutive MDA rounds, and whether MDA is delivered with increased frequency or coverage when treatment can recommence.

Supplementary Information A: Supplementary Methods provides a detailed description of the models. All (therapeutic) coverage levels refer to the percentage of people treated out of the total population, where total population includes children <5 years old and individuals who never take treatment (non-participation). Fig. S1 illustrates how elimination probabilities are calculated.

We followed the five principles of the Neglected Tropical Disease Modelling Consortium,^14^ to advocate and adhere to using good practice for policy-relevant modelling. Supplementary Table S1 presents the PRIME-NTD (Policy-Relevant Items for Reporting Models in Epidemiology of Neglected Tropical Diseases) table.

## Scenarios

### Treatment histories and transmission settings

We simulated a range of scenarios reflecting pre-control endemicity and historical treatment durations in the former Onchocerciasis Control Programme in West Africa (OCP, 1975–2002), specifically areas in the western extension in which ivermectin MDA was implemented without vector control,^15^ and the African Programme for Onchocerciasis Control (APOC, 1995–2015).^16^

### Pre-control endemicity

For the sake of completeness, we considered pre-intervention onchocerciasis endemicity levels given by baseline *Onchocerca volvulus* microfilarial prevalence of 20–85% (i.e. from hypoendemic to highly hyperendemic settings), although APOC prioritised treatment only in areas with microfilarial prevalence ≥40%.

### History of ivermectin MDA

Annual ivermectin MDA programmes were simulated with start years 2000, 2003, 2006, 2009, 2012, 2014, 2017 and 2020, assuming that 65% of the total population is treated per round (i.e. approximately 80% coverage of eligible individuals aged ≥5 years). The level of systematic non-participation was assumed to be 5%.

### Pre-existing biannual MDA programmes

We also modelled scenarios for pre-existing biannual programmes preceded by low coverage annual MDA motivated by the situation in the Madi-Mid North focus in Uganda,^17^ where the main vector is *Simulium damnosum sensu stricto* (for which both models are parameterised). These pre-existing biannual programmes were assumed to have started in 1994 with annual MDA at 25% coverage (due for instance, to internal conflict, as was the case of the Madi-Mid North focus) until 2012, when biannual treatment with 75% coverage of the total population (approximately 90% coverage of eligible individuals) began.^17^ A pre-intervention 50% microfilarial prevalence was motivated by the baseline values for Adjumani-Moyo,^16^ and Kitgum^18^ in the Madi-Mid North focus.

### Previous Ebola (2013–2014) outbreak

We modelled settings previously affected by the Ebola outbreak in western Africa in 2014, which were broadly motivated by the treatment histories in Sierra Leone, Liberia and Guinea.^19–21^ We assumed annual treatment from 2003 to 2013 (with coverage gradually increasing from 25% in 2003 to 65% in 2013), no treatment in 2014, 30% coverage in 2015, 65% coverage in 2016–2019, no treatment in 2020, 50% coverage in 2021, and 65% coverage from 2022–2030.

### Modelling interruptions due to COVID-19

We projected microfilarial dynamics and estimated elimination probabilities assuming either a 1-year (2020) or 2-year (2020 and 2021) interruption to ivermectin MDA. Additionally, we tested various assumptions of delays in achieving pre-COVID-19 coverage upon treatment resumption, e.g. missing MDA in 2020 but gradually increasing coverage from 30% in 2021, to 50% in 2022, and to 65% in 2023–2030. Levels of systematic non-participation were set at 5%.

### Modelling remedial strategies

We modelled the effectiveness of increasing frequency (to biannual treatment at 65% coverage) or therapeutic coverage (to 75% for annual treatment) as strategies for mitigating the impact of missed treatment rounds (either in 2020 or in 2020 and 2021). For a selection of the scenarios described above, we investigated the capacity for remedial treatment to revert the microfilaridermia trends and restore elimination probabilities (with MDA ceasing in 2030) to those predicted without interruption. Typically, MDP only approves ivermectin donations for biannual treatment if programmes can demonstrate sufficiently high coverage in preceding years. Therefore, we assumed that remedial biannual MDA followed only after a year of annual 65% coverage once programmes restart.

### Outcome measures

We present three model outputs to understand the implications of interruptions to MDA: (i) temporal trends in microfilarial prevalence (percent) to understand the short-term implications; (ii) probabilities of achieving elimination by 2030 to understand the long-term implications, and (iii) age-profiles for microfilarial prevalence (percent) and intensity (microfilariae/skin snip) to identify the most affected age groups.

## Results

### Impact of interruptions to ivermectin MDA for annual programmes

#### Temporal microfilarial prevalence trends

Fig. 1 shows, for EPIONCHO-IBM and ONCHOSIM, the microfilarial prevalence dynamics for programmes with a long (starting in 2000) and short (starting in 2017) history of ivermectin MDA, comparing no interruption with no treatment during 2020 (and a 30% coverage at resumption in 2021) and with no treatment during 2020 and 2021 (with a 30% coverage at resumption in 2022). The models qualitatively agree on how the temporal microfilarial prevalence trends during an interruption differ from those for continuous MDA through 2020–2021 for MDA starting in 2017. Increases in microfilarial prevalence after a 2-year interruption (2020–2021) are more pronounced than after a 1-year (2020) interruption. How coverage, once treatment recommences, influences the temporal dynamics of microfilarial prevalence after a 1-year interruption is shown in Supplementary Information B: Supplementary Results, Fig. S2 (EPIONCHO-IBM), and Fig. S3 (ONCHOSIM). A detailed discussion of how the dynamics following an interruption differ from those from without an interruption is presented in Supplementary Fig. S4.

**Figure 1.**
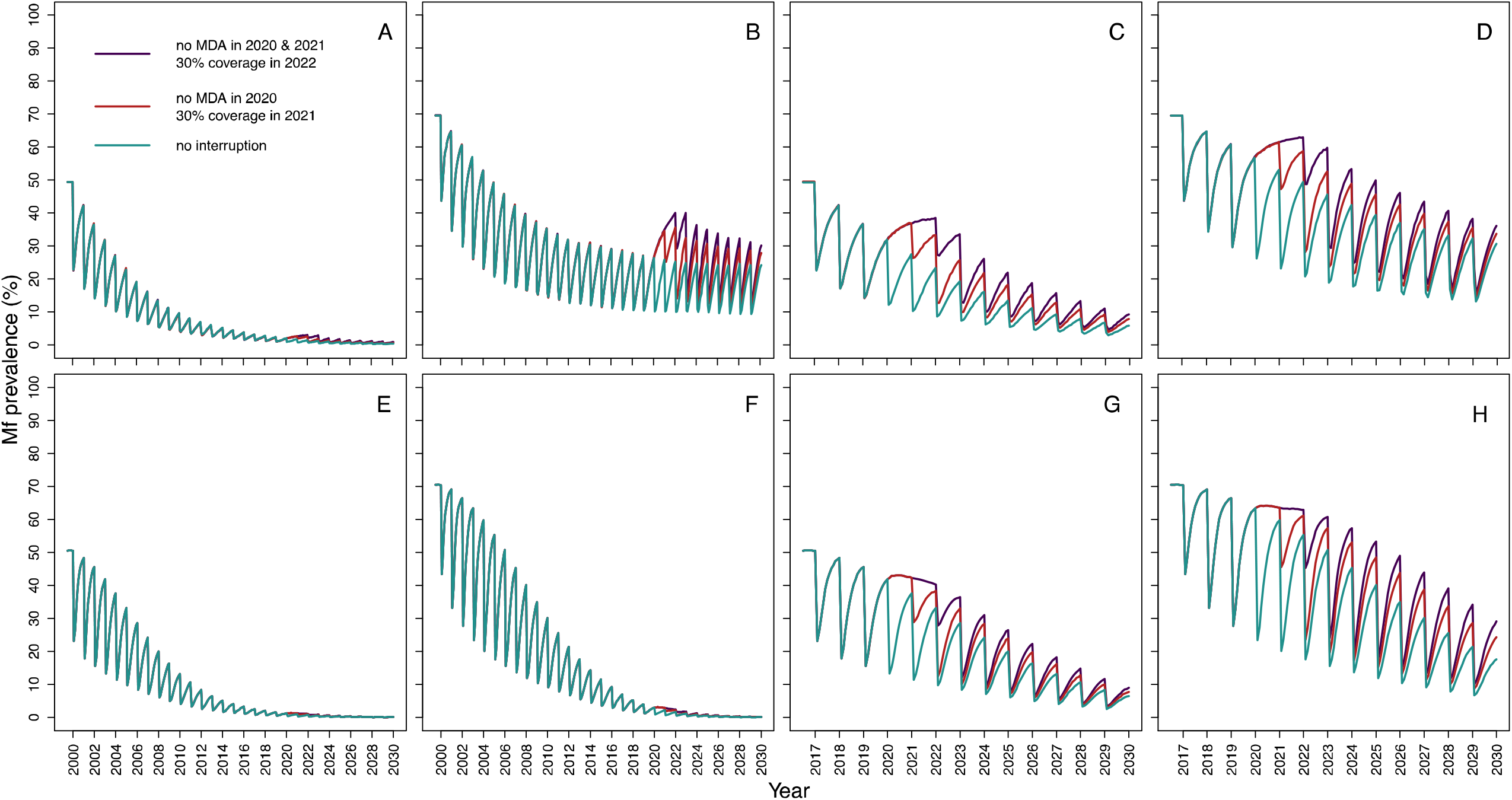
Temporal dynamics of *Onchocerca volvulus* microfilarial (Mf) prevalence assuming a 1-year (2020) and 2-year (2020–2021) interruption to annual mass drug administration (MDA) with ivermectin due to COVID-19 and no mitigation strategies, predicted by EPIONCHO-IBM (A-D) and ONCHOSIM (E-H). The pre-intervention (baseline) microfilarial prevalence in individuals aged ≥5 years is 50% (**A, C, E, G**), and 70% (**B, D, F, H**). Annual MDA occurs from 2000 to 2030 (early-start programmes, **A, B, E, F**) or from 2017 to 2030 (late-start programmes, **C, D, G, H**), assuming no treatment in 2020 only (red lines), no treatment in both 2020 and 2021 (violet lines), and no remedial strategies subsequently. The temporal microfilarial dynamics with no interruption to MDA are shown as green lines. The therapeutic coverage (of total population) is assumed to be 65%, with the exception of 30% in 2021 following a 1-year interruption and 30% in 2022 following a 2-year interruption to MDA. The proportion of systematic non-participation is set to 5% throughout all simulations.

### Elimination probabilities

Although EPIONCHO-IBM generally predicts lower elimination probabilities than ONCHOSIM, the models agree qualitatively on the impact of MDA interruptions due to COVID-19. Both models predict that interruptions to MDA will reduce the prospects of onchocerciasis elimination by 2030 if no mitigation strategies were implemented. Both models also predict that programmes with shorter treatment histories will be more vulnerable to MDA disruptions, than those which have distributed MDA for longer, particularly if treatment is delayed for two years (Fig. 2). The impact of a 1- and 2-year interruption on elimination probabilities with start years ranging from 2000 to 2020 is presented in Supplementary Fig. S5 and S6.

**Figure 2.**
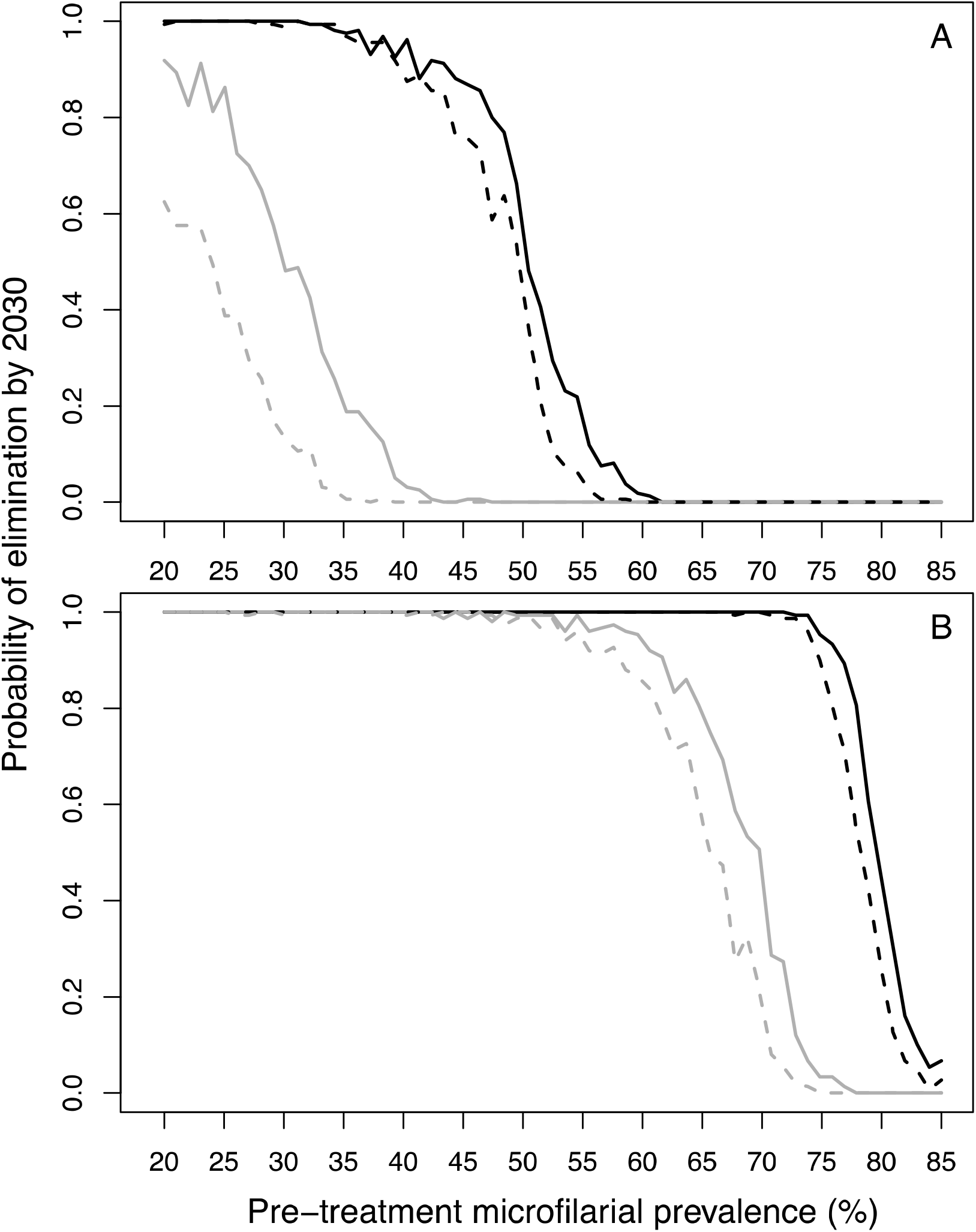
Elimination probabilities versus pre-intervention (baseline) endemicity (pre-treatment microfilarial prevalence) predicted by (A) EPIONCHO-IBM and (B) ONCHOSIM for early-start and late-start annual mass drug administration (MDA) with ivermectin with and without a 2-year (2020 and 2021) MDA interruption due to COVID-19 and no mitigation strategies. Annual MDA programmes start in 2000 (early-start, black lines) or 2017 (late-start, grey lines) and finish in 2030. Although in the African Programme for Onchocerciasis Control MDA treatment was prioritised for areas with microfilarial prevalence ≥40%, the range of baseline microfilarial prevalence in individuals aged ≥5 years explored is 20% – 85% for the sake of completeness. No interruption to MDA is represented by solid lines; a 2-year interruption is represented by dashed lines. The therapeutic coverage (of total population) is assumed to be 65%, with the exception of 30% in 2022. The proportion of systematic non-participation is set to 5% throughout all simulations.

### Mitigation strategies for annual programmes

#### Temporal microfilarial prevalence trends

Both models suggest that for a programme with a short treatment history, increasing treatment frequency, i.e. implementing remedial biannual MDA at 65% coverage upon resumption of MDA, will be more effective at reducing the impact of treatment disruptions than increasing treatment coverage (i.e. implementing remedial 75% coverage of annual MDA). We illustrate these results after a 2-year interruption (2020–2021) when MDA initially resumes at low annual coverage (30%, 2022), returns to annual 65% coverage (2023), and then either increases to a 6-monthly frequency for two consecutive years (2024–2025) at 65% coverage (Fig. 3), or increases to 75% annual MDA coverage also for two consecutive years (2024–2025) (Fig. 4). Remedial biannual treatment results in lower microfilarial prevalence leading up to 2030 (Fig. 4) than remedial increased coverage (Fig. 3). The temporal microfilarial prevalence dynamics for a 1-year interruption with either remedial increased treatment frequency or coverage are shown in Supplementary Fig. S7 and S8.

**Figure 3.**
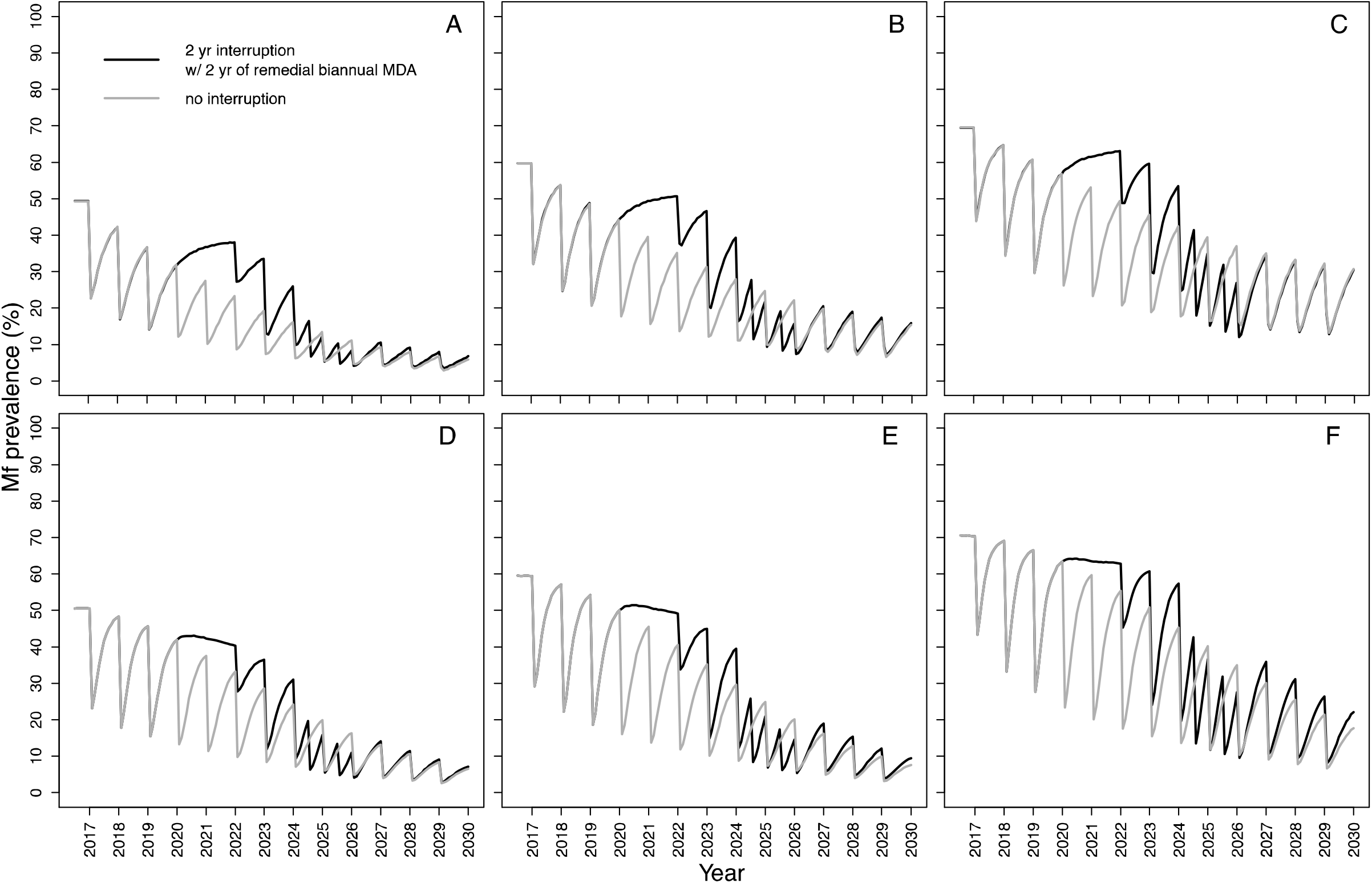
Temporal dynamics of *Onchocerca volvulus* microfilarial (Mf) prevalence predicted by EPIONCHO-IBM (A – C) and ONCHOSIM (D – F) with a 2-year (2020 and 2021) interruption due to COVID-19 of mass drug administration (MDA) with ivermectin (2017 – 2030) and remedial biannual MDA. The pre-intervention (baseline) microfilarial prevalence (in individuals aged ≥5 years) is: 50% (**A, D**), 60% (**B, E**), and 70% (**C, F**). Grey lines represent no interruption to MDA; black lines indicate no treatment in 2020 and 2021 but with two years of remedial biannual MDA in 2024 and 2025. The therapeutic coverage (of total population) is assumed to be 65%, with the exception of 30% in 2022. The proportion of systematic non-participation is set to 5% throughout all simulations.

**Figure 4.**
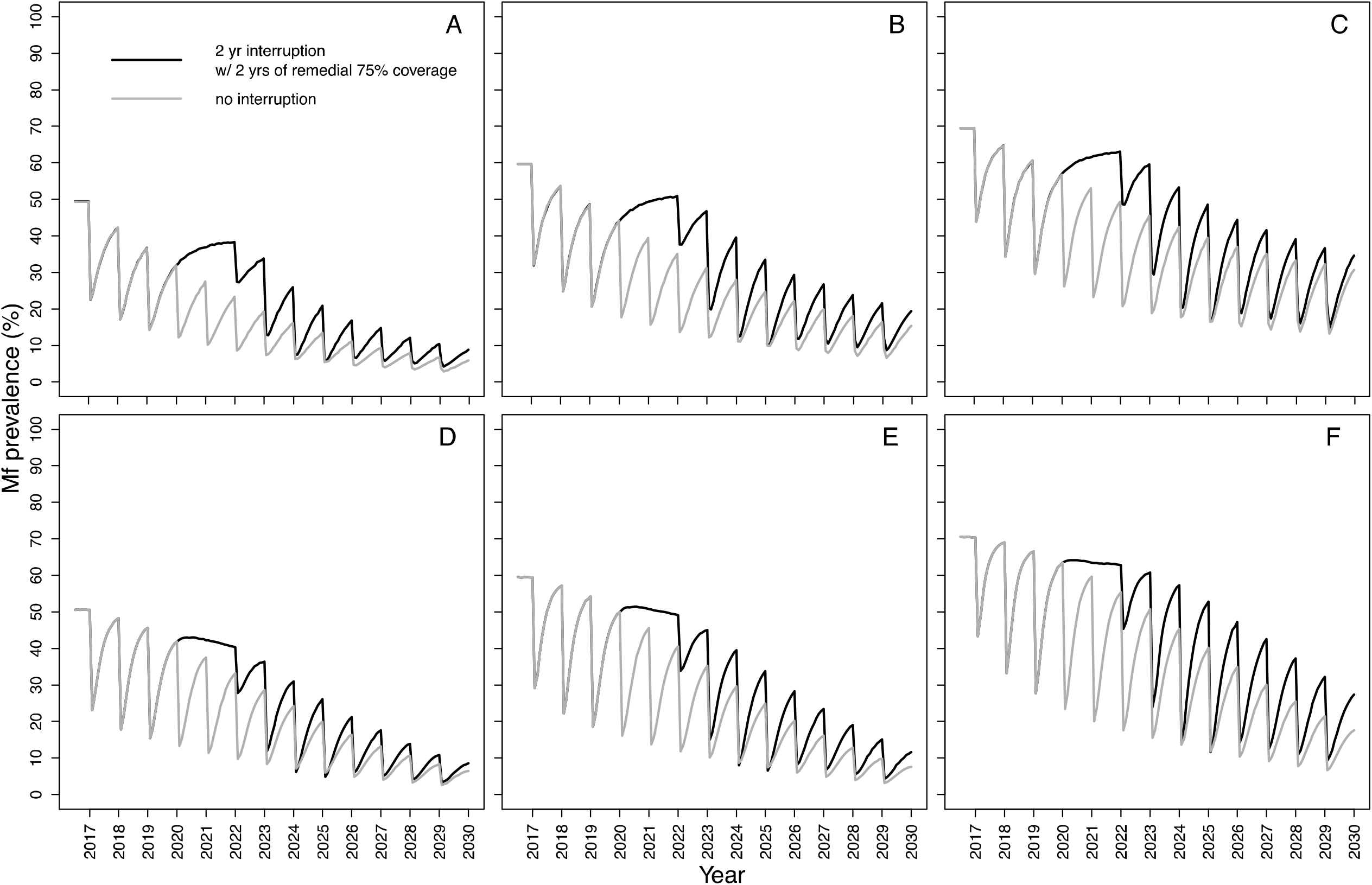
Temporal dynamics of *Onchocerca volvulus* microfilarial (Mf) prevalence predicted by EPIONCHO-IBM (A–C) and ONCHOSIM (D–F) with a 2-year (2020 and 2021) interruption due to COVID-19 of annual mass drug administration (MDA) with ivermectin (2017–2030) and remedial annual high-coverage MDA. The pre-intervention (baseline) microfilarial prevalence (in individuals aged ≥5 years) is: 50% (**A, D**), 60% (**B, E**), and 70% (**C, F**). Grey lines represent no interruption to MDA; black lines indicate no treatment in 2020 and 2021 but with two years of remedial high-coverage MDA in 2024 and 2025. The therapeutic coverage (of total population) is assumed to be 65%, with the exception of 30% in 2022, 75% in 2024 and 75% in 2025. The proportion of systematic non-participation is set to 5% throughout all simulations.

### Elimination probabilities with remedial strategies

For a 2-year interruption, although neither remedial strategy is sufficient to achieve the same elimination probabilities found in the absence of an interruption, 2 years of remedial biannual MDA results in higher elimination probabilities than remedial increase in coverage (Supplementary Fig. S9). For some treatment histories and pre-intervention endemicities (which do not result in 100% elimination probability without interruption), a 1-year of missed MDA followed by 1 year of remedial biannual MDA gives similar elimination probabilities to those in the absence of MDA disruptions (Supplementary Fig. S10).

### Pre-existing biannual programmes and those previously affected by Ebola

For the pre-existing biannual scenarios explored (which had achieved high pre-COVID-19 therapeutic coverage), a 1-year interruption did not result in pronounced microfilarial resurgence according to either model (Fig. 5). For this scenario, it was considered that programmes would resume treatment in 2021 given their strong performance before COVID-19, so a 2-year interruption was not investigated.

**Figure 5.**
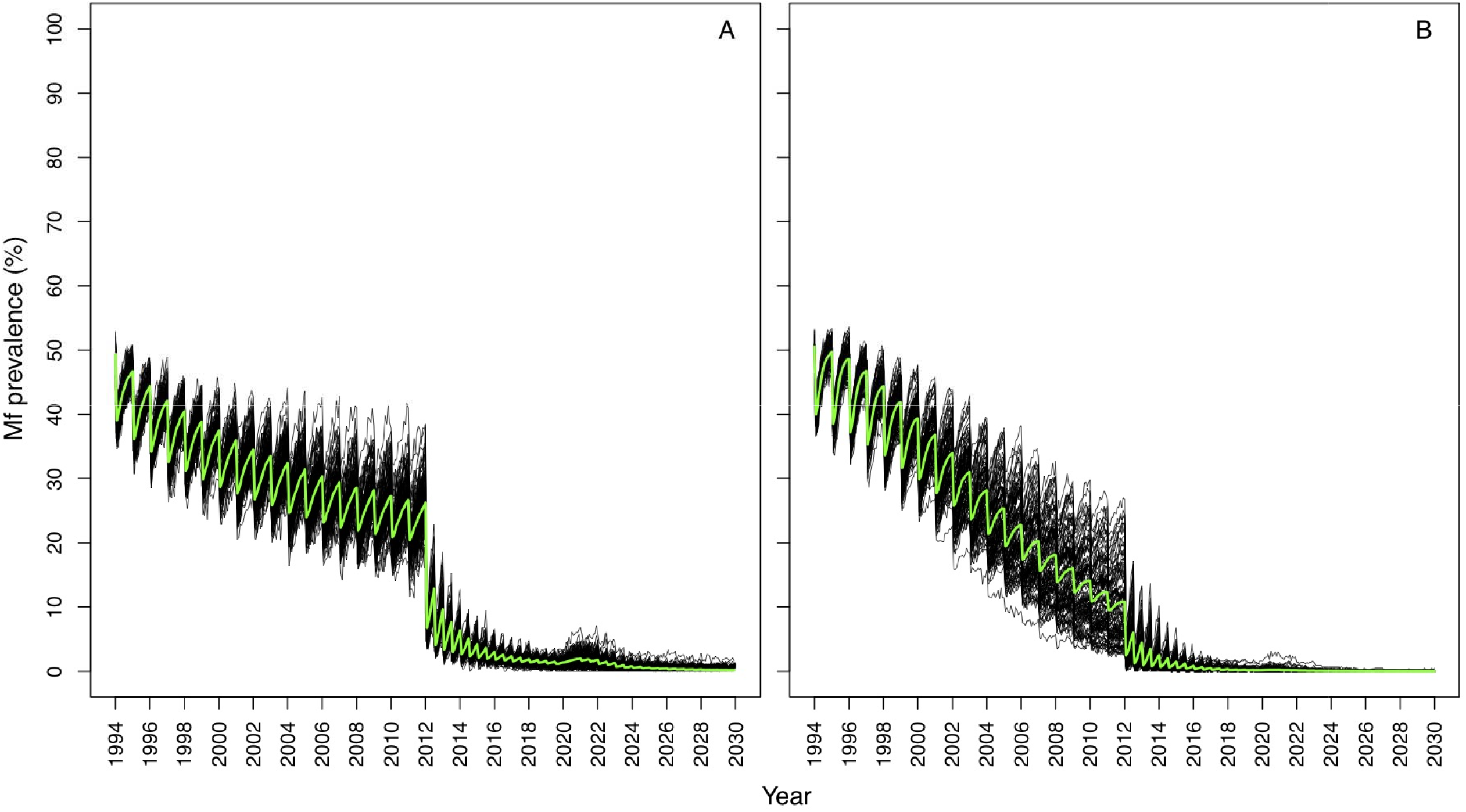
Temporal dynamics of *Onchocerca volvulus* microfilarial (Mf) prevalence predicted by EPIONCHO-IBM (A) and ONCHOSIM (B) in programmes with pre-existing biannual mass drug administration (MDA) with ivermectin and a 1-year interruption (2020) due to COVID-19. Annual MDA assumes low (25%) therapeutic coverage (of total population) from 1994 to 2011 and high (75%) biannual coverage (of total population) from 2012 to 2019 (motivated by the Madi-Mid North focus in Uganda, where the main vector is *Simulium damnosum sensu stricto*), with a 50% pre-intervention (baseline) microfilarial prevalence (in individuals aged ≥5 years). Following resumption of MDA in 2021, coverage (of total population) is assumed to reach 75% (same as pre-interruption). Black lines indicate individual simulations. Green lines indicate the mean of all simulations. The proportion of systematic non-participation is set to 5% throughout all simulations.

EPIONCHO-IBM predicts that the impact of a 1-year interruption due to COVID-19 in 2020 on microfilarial prevalence dynamics in West African programmes that were affected by the Ebola outbreak in 2014 will depend (like in other scenarios) on transmission setting (Fig. 6). When assuming a 50% baseline microfilarial prevalence, there were small differences between no interruption and a 1-year interruption by 2030; however, for the 70% baseline microfilarial prevalence settings, clear differences between no interruption and a 1-year interruption were still evident by 2030 according to both models.

**Figure 6.**
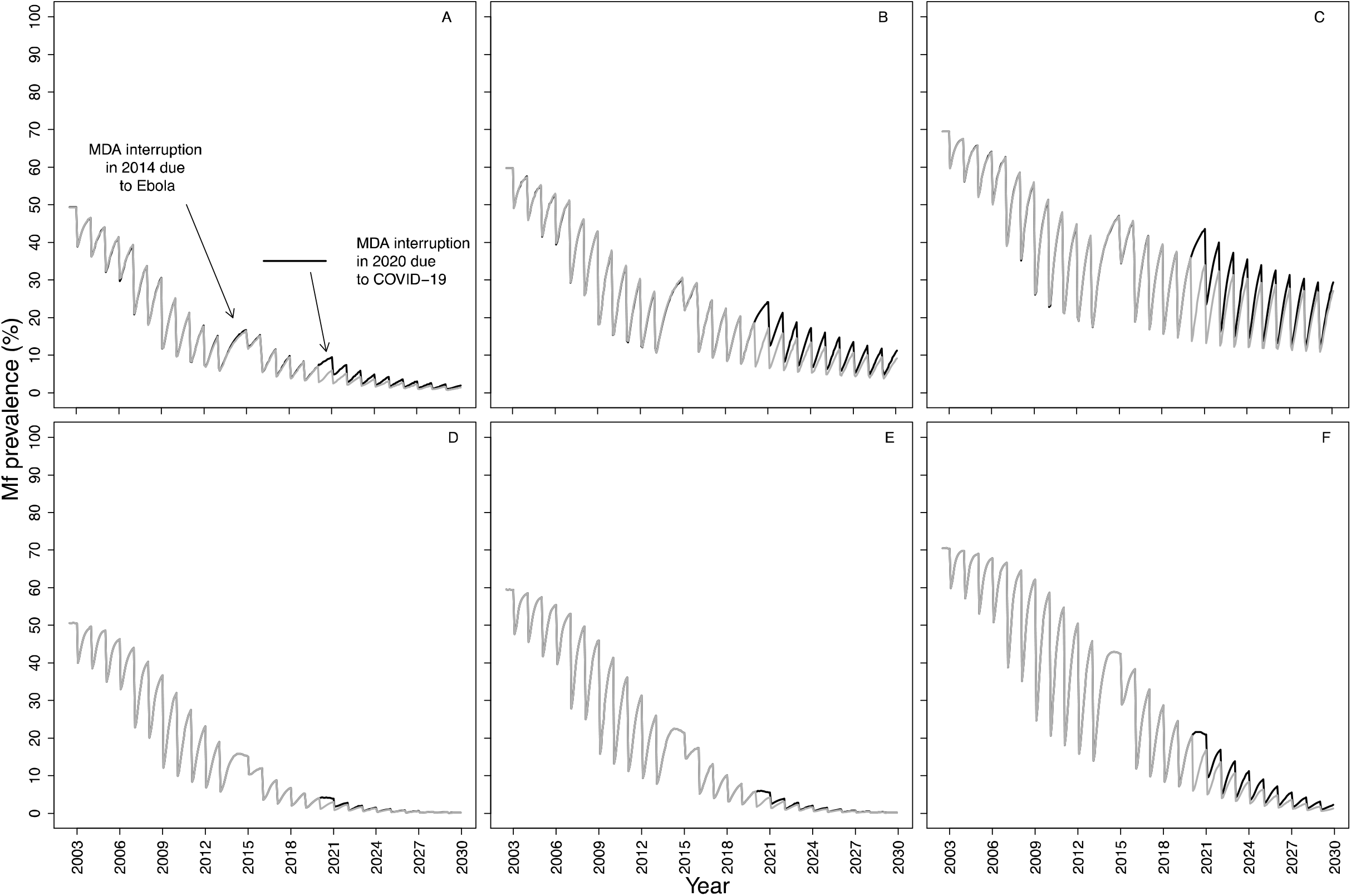
Temporal dynamics of *Onchocerca volvulus* microfilarial (Mf) prevalence predicted by EPIONCHO-IBM (A, B, C) and ONCHOSIM (D, E, F) during annual mass drug administration (MDA) with ivermectin, assuming a 1-year interruption (2014) due to the Ebola epidemic in West Africa, a 1-year interruption due to COVID-19 in 2020 and no remedial strategies. The pre-intervention (baseline) microfilarial prevalence in individuals aged ≥5 years is 50% (**A, D**), 60% (**B, E**) and 70% (**C, F**). The therapeutic coverage (of total population) is assumed to be 25% in 2003 and 2004, 30% in 2005 and 2006, 50% in 2007 and 2008, 65% in 2009 and 2012, 30% in 2015 and 65% from 2016 onwards (motivated by the situation in West African countries that had experienced civil conflict and were affected by the Ebola epidemic), with the exception of 2021 (50%). Black and grey lines indicate, respectively, the microfilarial dynamics with and without an interruption due to COVID-19. The proportion of systematic non-participation is set to 5% throughout all simulations.

### Age profiles of microfilarial prevalence and intensity

EPIONCHO-IBM and ONCHOSIM predict noticeably different age (and sex) patterns of microfilarial prevalence and intensity after three rounds of annual ivermectin MDA, with or without a 2-year (2020–2021) interruption (with MDA starting in 2017 and initial microfilarial prevalence of 70%) (Fig. 7). EPIONCHO-IBM predicts higher prevalence and intensity in children under the age of ten years following a 2-year MDA interruption than when there is no MDA interruption, and this is particularly pronounced in boys (Fig. 7A and 7B for prevalence; 7E and 7F for intensity). Following the interruption, ONCHOSIM predicts very low infection prevalence and intensity in children under 5 years, and an evident, but smaller increase in 5–10-year olds than in EPIONCHO-IBM (Fig. 7C and 7D for prevalence; 7G and 7H for intensity). By contrast, ONCHOSIM predicts a marked microfilarial prevalence and intensity increase in the ≥20 years olds compared to EPIONCHO-IBM. (The baseline profiles are shown as insets in Fig. 7; individual-level microfilarial load is illustrated in Supplementary Fig. S11.)

**Figure 7:**
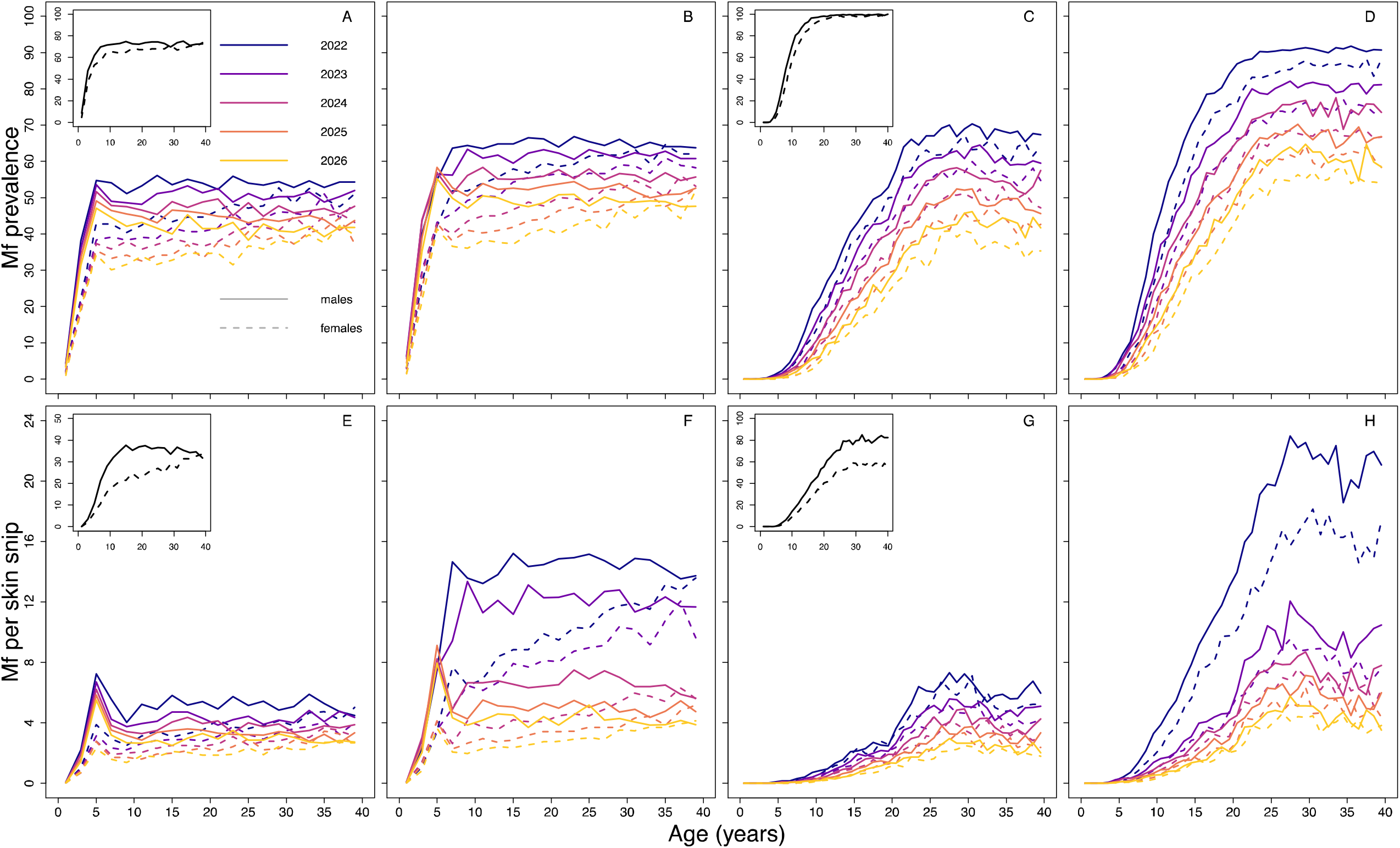
Age- and sex-profiles of *Onchocerca volvulus* microfilarial (Mf) prevalence (percent, A – D) and mean intensity (mf/skin snip, E – H) predicted by EPIONCHO-IBM (A, B, E, F) and ONCHOSIM (C, D, G, H) assuming no interruption to mass drug administration (MDA) with ivermectin (A, C, E, G) and a 2-year interruption to MDA in 2020 and 2021 (B, D, F, H) due to COVID-19. The pre-intervention (baseline) microfilarial prevalence is 70% in individuals aged ≥5 years and the age-profiles for prevalence (**A, C**) and intensity (**E, G**) at baseline are shown in the figure insets. Annual MDA starts in 2017 and the profiles (solid lines for males and dashed lines for females) are shown for 2022 (black), 2023 (violet), 2024 (purple), 2025 (red) and 2026 (yellow). The therapeutic coverage (of total population) is assumed to be 65%, with the exception of 30% in 2022 (**B, D, F, H**). The proportion of systematic non-participation is set to 5% throughout all simulations.

## Discussion

While in the grip of the COVID-19 pandemic in April 2020, the WHO recommended to suspend all epidemiological surveys and MDA activities for NTDs to help curtail the transmission and spread of the SARS-CoV-2 virus.^1^ These disruptions to ivermectin MDA created concern in the NTD community regarding potential short-term increases in transmission and impacts on longer-term elimination prospects for onchocerciasis, particularly in the context of the EOT goals proposed by the WHO in its 2021–2030 NTD roadmap.^5^ In response to these concerns, we have used two stochastic onchocerciasis transmission models, EPIONCHO-IBM and ONCHOSIM, to investigate the impact of (1- and 2-year) interruptions to MDA in a variety of settings motivated by the epidemiological situation in endemic areas in Africa.

Generally, the models suggest that programmes with short treatment histories of pre-existing annual MDA (late-start programmes) will be the most vulnerable to a two-year (2020–2021) interruption. Programmes with longer treatment histories (early-start programmes) could also be adversely affected if initial endemicity levels indicate intense (hyperendemic) transmission. The influence of baseline microfilarial prevalence was more pronounced in EPIONCHO-IBM for both early- and late-start MDA programmes than in ONCHOSIM. Although for late-start programmes a 1-year interruption to MDA impacted the elimination probabilities and microfilarial prevalence in 2030, the effect would be more tolerable than for a 2-year interruption; early-start programmes with lower pre-intervention endemicities were mostly unaffected.

In July 2020, the WHO issued guidelines for the resumption of MDA provided it can be delivered ‘safely’ following a case-by-case risk-benefit assessment, with due consideration of a health system’s capacity to conduct such modified activities effectively in the context of the ongoing COVID-19 pandemic.^2^ Therefore, the implementation of mitigation strategies in the models allowed for a gradual scaling-up of therapeutic coverage once MDA resumes. Both models predict that remedial biannual MDA (in 2024 and 2025, after having demonstrated up-scaling to 65% coverage through 2022 and 2023) would be more effective at controlling increases in infection and decreases in elimination probabilities than remedial high (75%) coverage of annual MDA (also in 2024 and 2025).

Both models also predicted that pre-existing biannual MDA programmes—which had achieved a high treatment coverage for nearly a decade (in our simulations 75% pre-COVID-19 coverage from 2012 inclusive to reflect a 90% coverage of eligible population)—would not be adversely affected by a 1-year interruption. This was despite assuming a slow start of annual MDA at low coverage (before implementing biannual MDA) to capture initial difficulties in implementing treatment.^17^ A 1-year (rather than a 2-year) interruption was explored for these settings because of their pre-COVID-19 strong programmatic performance.^17^ It was also assumed that these programmes would be able to resume biannual MDA at the same high coverage they had recorded before the pandemic given their existing structure and commitment to achieve EOT.

Some West African countries (e.g. Guinea, Liberia, Sierra Leone) had already had their ivermectin MDA programmes interrupted in 2014 due to the Ebola epidemic. We therefore considered this situation, in addition to a 1-year COVID-19 interruption in 2020. Both models predicted that in mesoendemic settings (50% baseline microfilarial prevalence), these two separate 1-year interruptions would not greatly impact on programmes’ performance, provided that coverage levels can catch up reasonably quickly following resumption of MDA (50% in 2021 and 65% in 2022 onwards). In hyperendemic and highly hyperendemic settings (60% and 70% baseline microfilarial prevalence) the impact would be more pronounced (particularly according to EPIONCHO-IBM).

Although programmes measure progress in terms of reducing mean infection prevalence and intensity at population level, favourable infection trends may mask heterogeneities among different population sub-groups. In areas of intense transmission and only recent MDA implementation, which could be exemplified by the situation of the Maridi Dam in South Sudan,^22^ children may reach their fifth birthday (when they become eligible for ivermectin treatment) with a sizeable microfilarial load. This has been shown to be a significant risk factor for the development of epilepsy later in life,^9^ and could also contribute to increasing the relative risk of mortality, which for a given microfilarial load is higher in children than in adults.^23^ In the Maridi villages, Ov-16 serology (a marker of exposure to infection) revealed a 20% (95% CI = 13%–29%) seroprevalence in the 3–6-year-olds.^22^ Following a 2-year MDA interruption, children may not receive treatment until their 7^th^ birthday, leading to a further microfilarial load build-up. Our modelling results indicate that in late-start (e.g. 2017) and high initial endemicity programmes (e.g. 70% microfilarial prevalence), there might be substantial increases in infection intensity in children aged <10 years, particularly according to EPIONCHO-IBM. In addition, both models predict that in these settings, and following a 2-year MDA interruption, there will also be increases in infection prevalence and intensity in older age groups (moderate in EPIONCHO-IBM but pronounced in ONCHOSIM). This increase could lead to exacerbation of other onchocerciasis-associated sequelae, such as troublesome itch, according to epidemiological^8^ and modelling studies linking infection and disease.^24,25^ These results are mostly determined by the assumed patterns of age- and sex-dependent exposure to infection, which differ markedly between the two models.^26^

Although the models agree qualitatively on the role of treatment history in the outcome of interruptions to ivermectin MDA, and the differences between remedial strategies in mitigating the ensuing setbacks, ONCHOSIM predicts higher elimination probabilities than EPIONCHO-IBM. This is partly because EPIONCHO-IBM assumes strong regulatory processes operating on parasite establishment within humans that are relaxed as transmission declines during interventions, increasing the stability and resilience of the host–parasite system; these processes are not assumed in ONCHOSIM.^12,26,27^ In addition, the aforementioned differences in the assumed age- and sex-specific exposure patterns, and associated age and sex profiles of infection results in ONCHOSIM predicting a lower microfilarial intensity in children under 5, while in EPIONCHO-IBM there exists a larger reservoir of infection in untreated children, which also contributes to lower elimination probabilities.

An assumption made in most of the modelled scenarios model, is that when MDA resumes coverage will initially be lower than pre-pandemic levels and could take several years to recover. However, the usual drug distribution modality, by which members of the community present at a focal point to receive ivermectin, could be replaced by door-to-door drug distribution (as a result of social distancing measures). Depending on local circumstances, some programmes may achieve well-documented and high coverage levels, although the latter would also rely on well-stocked supply chains.

We did not consider the potential use of moxidectin and/or vector control as alternative remedial strategies to mitigate the impact of the current pandemic. This is because, while these approaches are very promising,^28–30^ they are currently not operationally implemented by national control programmes. However, where vector control is practicable, it should be considered as a complementary intervention, when it is more conducive than chemotherapeutic approaches to implementation in a socially-distanced manner.^2^

## Conclusions and Recommendations

Both EPIONCHO-IBM and ONCHOSIM indicate that ivermectin MDA programmes with shorter treatment histories will be most vulnerable to MDA interruptions caused by the COVID-19 pandemic, particularly if treatment cannot be resumed safely in 2021. This impact may be noticeable in local infection resurgence in higher endemicity settings and may reduce the probability of achieving EOT by 2030. Programmes with longer treatment histories of annual MDA that have achieved and maintained a coverage of 65% are predicted to be less affected (but not totally impervious) to interruptions of ivermectin MDA, particularly if high initial endemicity indicates highly propitious transmission conditions. Young children have the potential to be negatively affected by increased levels of transmission resulting from missed MDA rounds, particularly for a 2-year interruption in highly endemic settings recently incorporated to ivermectin MDA programmes, depending on local age-exposure patterns.

The impact of COVID-19 on progress towards the WHO 2021–2030 goals is best ameliorated by implementing biannual MDA as soon as pre-COVID-19 levels of therapeutic coverage are restored (in our simulations to 65% of the total population in 2024–2025). This mitigation strategy is indicated as more effective than increasing annual MDA coverage (to 75%). MDA programmes should promptly conduct the risk-benefit evaluations indicated by the WHO,^2,31^ so alternative modalities of MDA delivery can be put in place safely and effectively to resume MDA and regain pre-COVID-19 levels of coverage where possible (e.g. door-to-door distribution with household members measuring themselves with disinfected height poles and community drug distributors leaving tablets at the doorstep and observing household members swallowing the tablets with safe social distancing). Additionally, in August 2020, The United States Agency for International Development (USAID) released guidance on how best to implement MDA in the context of COVID-19.^32^ To complement the WHO and USAID documentation, we offer a decision-making tree to summarise our results and recommendations which we hope will help programme managers to navigate the landscape of ivermectin MDA for onchocerciasis during the ongoing pandemic (Fig. 8).

**Figure 8.**
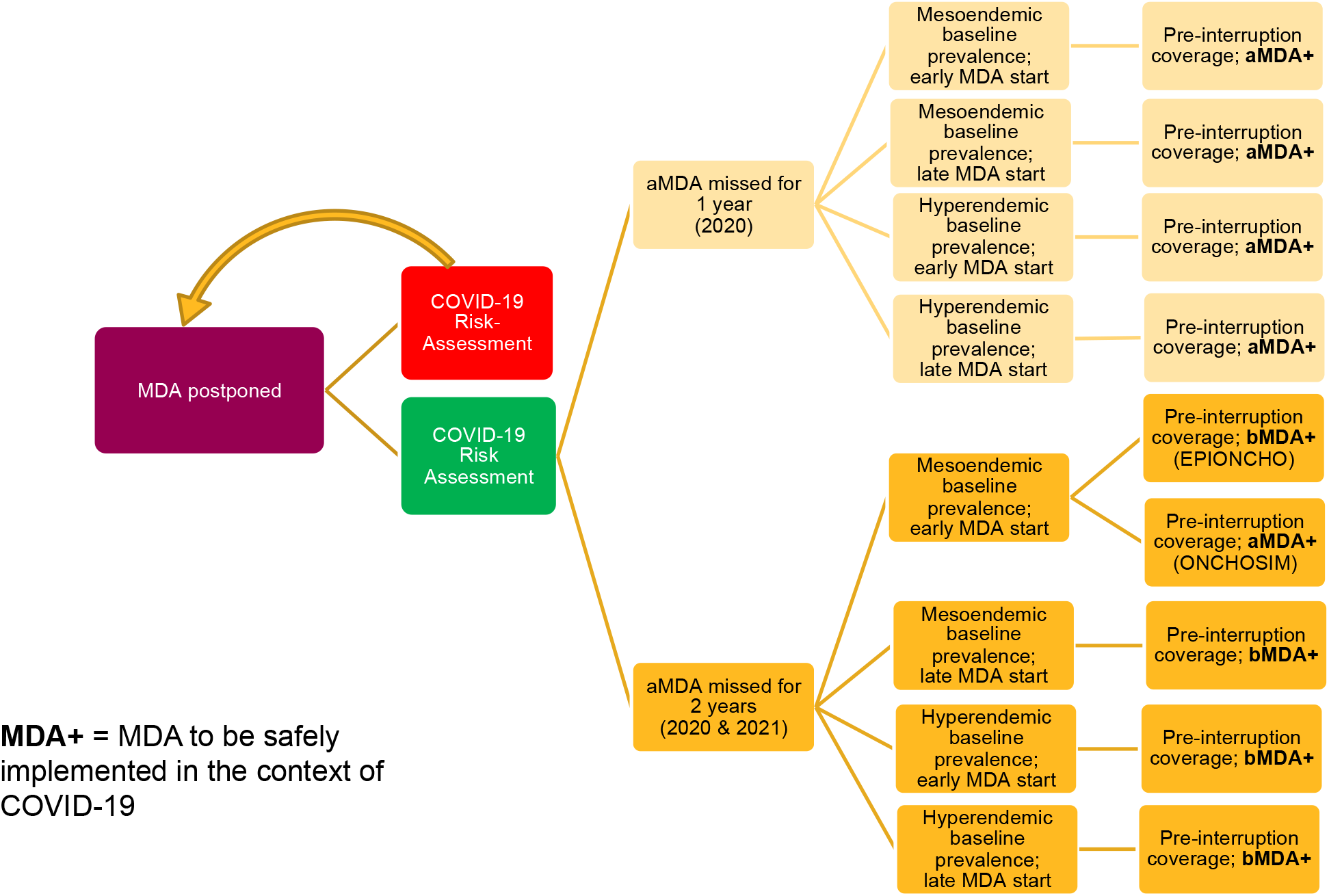
Decision tree for implementing ivermectin MDA following a 1- or 2-year interruption to treatment programmes as a result of COVID-19. COVID-19 risk assessment conducted but risk deemed too high to safely implement MDA is represented by the red box, in which case MDA is postponed until subsequent risk assessments allow treatment activities to recommence. COVID-19 risk assessment conducted and deemed that MDA can resume safely is represented by the green box. MDA strategy upon resumption is based on duration of interruption, baseline prevalence, and treatment history. MDA interruption (or missed MDA) is defined as no MDA in the period specified, i.e. 1 year (≥12 months since the last round of MDA) or 2 years (≥24 months since the last round of MDA). One year of missed annual MDA followed by a remedial biannual round in the following year can be interpreted as delayed MDA. Pre-interruption coverage assumes that before COVID-19 programmes had reached the minimum effective coverage of 65% of total population and that this value wil be reached following resumption of MDA. Early MDA start = programmes starting MDA in 2000; late MDA start = programmes commencing MDA in 2017 (these start times were selected as they are at the two extremes of the treatment durations simulated); aMDA = annual MDA; bMDA = biannual MDA; MDA+ = MDA implemented following World Health Organization guidelines for minimising the risk of COVID-19 transmission.^2,31^

## Data Availability

Data availability statement: No new data were used in this study. The model code is available on Github.

## Abbreviations

ABR: annual biting rate
APOC: African Programme for Onchocerciasis Control
EOT: elimination of transmission
IBM: individual-based model
MDA: mass drug administration
aMDA: annual mass drug administration
bMDA: biannual mass drug administration
MDP: Mectizan Donation Program
NTD: neglected tropical disease
OCP: Onchocerciasis Control Programme in West Africa
PCT: preventive chemotherapy and transmission control
USAID: United States Agency for International Development
WHO: World Health Organization.

## Authors contributions

JIDH: Conceptualization, Methodology, Investigation, Formal analysis, Software, Visualization, Writing – original draft, Writing – review & editing

DJB: Conceptualization, Formal analysis, Software, Writing – review & editing

MW: Conceptualization, Methodology, Supervision, Writing – review & editing

PM: Methodology, Writing – review & editing

ADH: Conceptualization, Writing – review & editing

LCH: Conceptualization, Writing – review & editing

PD: Visualization, Writing – review & editing

SJdV: Funding acquisition, Supervision, Writing – review & editing

WAS: Conceptualization, Supervision, Writing – review & editing

MGB: Conceptualization, Methodology, Funding acquisition, Supervision, Visualization, Writing – original draft, Writing – review & editing

## Code availability

The code of EPIONCHO-IBM is available at https://github.com/jonathanhamley/EPIONCHO-IBM. The code of ONCHOSIM is available at https://gitlab.com/erasmusmc-public-health/wormsim.previous.versions/-/blob/master/wormsim-2.58Ap27.zip

## Description of SI File

Detailed description of modelling methodology and additional results.

