## Supplementary information for "What does the COVID-19 pandemic mean for the next decade of onchocerciasis control and elimination?"

^a^ London Centre for Neglected Tropical Disease Research (LCNTDR), Department of Infectious Disease Epidemiology, School of Public Health, Faculty of Medicine (St Mary’s campus), Imperial College London, Norfolk Place, London W2 1PG, UK; ^b^ MRC Centre for Global Infectious Disease Analysis, Department of Infectious Disease Epidemiology, School of Public Health, Faculty of Medicine (St Mary’s campus), Imperial College London, Norfolk Place, London W2 1PG, UK; ^c^ Department of Public Health, Erasmus MC, University Medical Center Rotterdam, P.O. Box 2040, 3000 CA Rotterdam, The Netherlands.^d^ London Centre for Neglected Tropical Disease Research (LCNTDR), Department of Pathobiology and Population Sciences, Royal Veterinary College, University of London, Hatfield AL9 7TA, UK; ^e^ Neglected and Disabling Diseases of Poverty Consultant, Kent, UK; ^f^ Sightsavers, 35 Perrymount Road, Haywards Heath, RH16 3BW UK.

**A: Supplementary metho****ds**

***Mathematical modelling***

***Models***

We use two stochastic onchocerciasis transmission models, namely, EPIONCHO-IBM^1,2^ (see Walker et al. and Basáñez et al. for descriptions of deterministic versions)^3,4^ and ONCHOSIM.^5,6^ The pre-intervention (baseline) simulations (20%–85% microfilarial prevalence, at increments of 1% prevalence) were simulated by allowing the annual biting rate (ABR, the number of vector bites per person per year) to vary for EPIONCHO-IBM. For ONCHOSIM, in addition to the ABR, changing the level of heterogeneity in exposure to vector bites and including an external force of infection (required to stabilise low, endemic prevalence values) were also considered. The vectors of human onchocerciasis are blackflies of the genus *Simulium* (Diptera: Simuliidae), and both models are parameterised for the savannah species of the *Simulium damnosum sensu lato* (*s.l.*) complex (i.e., *Simulium damnosum sensu stricto* (s.s.)/*S. sirbanum*). These vector-specific parameterisations refer to vector competence (the proportion of ingested *Onchocerca volvulus* microfilariae that develop to infective, L3 larvae), parasite larval progression rates within the flies, proportion of blood-meals taken on humans, and vector survival. In both models, the parasite dynamics in the fly population are modelled deterministically.^3^ Due to the different relationship that exists between microfilarial prevalence and ABR in EPIONCHO and ONCHOSIM,^4^ the latter implements an external force of infection (a source of onchocerciasis infection external to that of the setting being considered in order to stabilise low prevalence). Individual baseline simulations for each prevalence bin were, therefore, based on the microfilarial prevalence (of a single simulation) at (pre-intervention) equilibrium rather than by ABR, which would have resulted in the mean of many simulations with the same ABR producing the target microfilarial prevalence.

***Elimination***

The models were run 200 times for each pre-intervention microfilarial prevalence and treatment scenario. These model runs varied in terms of the ABR (EPIONCHO-IBM) and ABR, external force of infection and individual-level exposure heterogeneity (ONCHOSIM), with each model run yielding the target microfilarial prevalence to cover the aforementioned range 20%–85% microfilarial prevalence, at increments of 1% prevalence. From these 200 models runs, the proportion in which the microfilarial prevalence is 0, 50 years after the last round of mass drug administration (MDA) with ivermectin in 2030, gives the elimination probability. This procedure is illustrated in Fig. S1 for EPIONCHO-IBM and ONCHOSIM, depicting the temporal dynamics of microfilarial prevalence, both during 30 years of annual MDA and a 2-year interruption (2020 and 2021) to ivermectin treatment (due to the disruptions to treatment programmes caused by the COVID-19 pandemic), and 10 years after the last round of MDA, to show resurgence or elimination. Although the mean microfilarial prevalence predicted by EPIONCHO-IBM during the interruption remains low, some of the individual simulations show a pronounced bounce back, illustrating the uncertainty in model predictions, and highlighting that although the temporal dynamics of infection prevalence predicted by both models are very similar, the rates of recrudescence can be very different. As outlined in the Discussion section of the Main Text, these differences are centred around contrasting assumptions of parasite establishment within humans in both models, and differences in age- and sex-specific exposure profiles.^7^

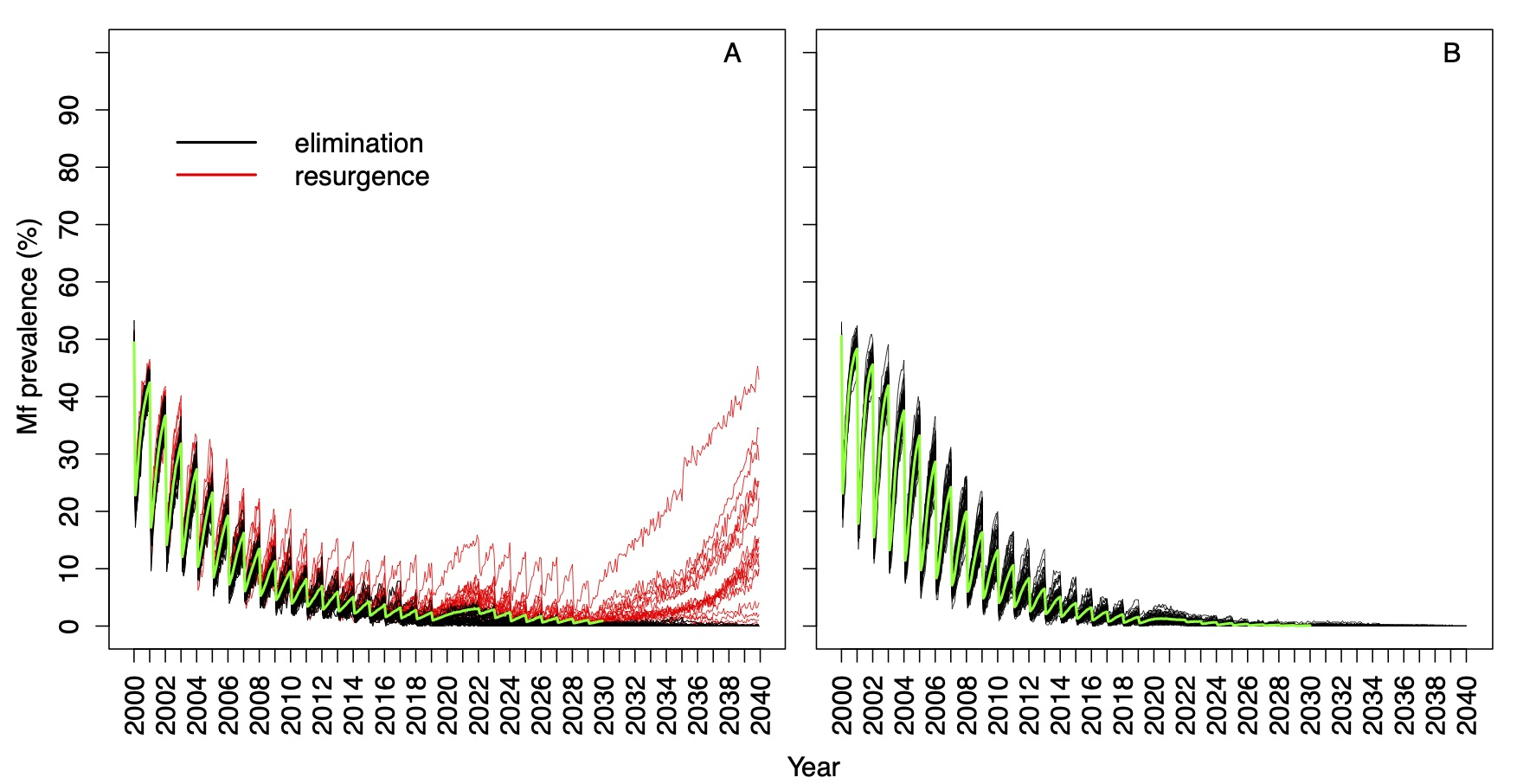

**Figure S1. Temporal dynamics of *Onchocerca volvulus* microfilarial (Mf) prevalence during 30 years of ivermectin MDA predicted by (A) EPIONCHO-IBM and (B) ONCHOSIM.** Dynamics are shown for individual (repeat) simulations in a setting with a 50% pre-intervention (baseline) microfilarial prevalence in individuals aged ≥5 years, with a 2-year interruption (2020 and 2021) to annual mass drug administration (MDA) with ivermectin due to COVID-19, and for 10 years after the last round of MDA in 2030. Black lines indicate simulations resulting in the elimination of *O. volvulus*; red lines indicate resurgence (no elimination). The green line represents the mean of both the black and red lines. Therapeutic coverage is assumed to be 65% of total population, with the proportion of systematic non-participation set to 5% in all simulations.

***Age- and sex-specific microfilarial prevalence and intensity profiles***

The scenarios used to produce the age- and sex-specific microfilarial prevalence and intensity profiles were run a total of 400 times for each model. The microfilarial intensity output in EPIONCHO-IBM is the mean number of microfilariae per mg of skin (mf/mg), whereas the corresponding output in ONCHOSIM is expressed as microfilariae per skin snip (mf/ss). Therefore, to make the intensity outcomes of both models comparable, it was assumed that one skin snip (taken with a 2-mm Holth corneo-scleral punch) weighs approximately 2 mg.^3^

***Therapeutic coverage and treatment non-adherence (non-participation)***

In both models, we define the (therapeutic) coverage as the percentage of the total population which receives treatment. Therefore, coverage can never be 100%, since a proportion of the population is younger than 5 years and therefore not eligible for treatment. In both models, we assume that a proportion of the population systematically does not participate (5% for all simulations and both models), i.e. they never receive treatment. Therefore, as the systematic non-participation increases, the upper limit for possible total population coverage decreases. Note that ONCHOSIM assumes a sex- and age-dependent treatment adherence,^5^ which is not present in EPIONCHO-IBM.

***Adherence to the five principles of the NTD Modelling Consortium***

We demonstrate how we followed the five principles of the Neglected Tropical Disease (NTD) Modelling Consortium^8^ by completing the ‘PRIME-NTD’ table (Policy-Relevant Items for Reporting Models in Epidemiology of Neglected Tropical Diseases) in Table S1 below.

**Table S1. PRIME-NTD (Policy-Relevant Items for Reporting Models in Epidemiology of Neglected Tropical Diseases) Summary Table^8^**

| Principle | What has been done to  satisfy the principle? | Where in the manuscript is this described? |
| --- | --- | --- |
| 1. **Stakeholder engagement** | Three authors of the paper are or have been involved in large-scale treatment programmes and played a central role in the selection of the simulated scenarios. LCH is a Senior Technical Advisor at Sightsavers, PD is the Technical Director for NTDs at Sightsavers, and ADH has been involved in delivering primary health care and ophthalmic services for NTDs for around 45 years. He was also the former director of the Mectizan Donation Programme. | Author list and author contributions. |
| 2. **Complete model documentation** | References have been included which contain full documentation for the EPIONCHO-IBM and ONCHOSIM models, and links to repositories for model code have been provided. | In the Main Text references, Supplementary Information and under the ‘Code availability’ section in the Main Text. |
| 3. **Complete description of data used** | No actual data were used. When the epidemiological scenarios simulated were motivated by particular examples (e.g. pre-existing biannual programmes; programmes formerly affected by Ebola), references have been provided in the Main Text to document this motivation. | NA |
| 4. **Communicating uncertainty** | We account for uncertainty in the parameters which determine baseline microfilarial prevalence (e.g. annual biting rate, heterogeneity in exposure to vector bites). Structural uncertainty is accounted for by making predictions with two models that differ in their structural assumptions regarding regulation of infection processes in humans and age- and sex-specific exposure patterns).^7^ | In Supplementary Information A: Supplementary methods. |
| 5. **Testable model outcomes** | The temporal infection trends and age profiles for infection predicted by the models have the potential to be tested empirically. | In the Results section. |

**B: Supplementary results**

***Influence of therapeutic coverage following ivermectin MDA resumption***

How therapeutic coverage, once treatment recommences, influences the temporal dynamics of microfilarial prevalence after a 1-year (2020) interruption to ivermectin MDA due to COVID-19 is shown in Fig. S2 (EPIONCHO-IBM), and Fig. S3 (ONCHOSIM) below. The green lines assume that coverage (of total population) in 2021 is 30%, followed by 50% in 2022 and 65% in 2023. The violet lines assume that coverage in 2021 is 30% followed by 65% in 2022 and 2023. The blue lines assume that the coverage in 2021, 2022 and 2023 is 65%, same as in the pre-COVID-19 period. From 2024 onwards, assumed therapeutic coverage is 65%. In all simulations the proportion of systematic non-participation is set to 5%, and it is assumed that this does not change throughout the period that MDA is delivered.

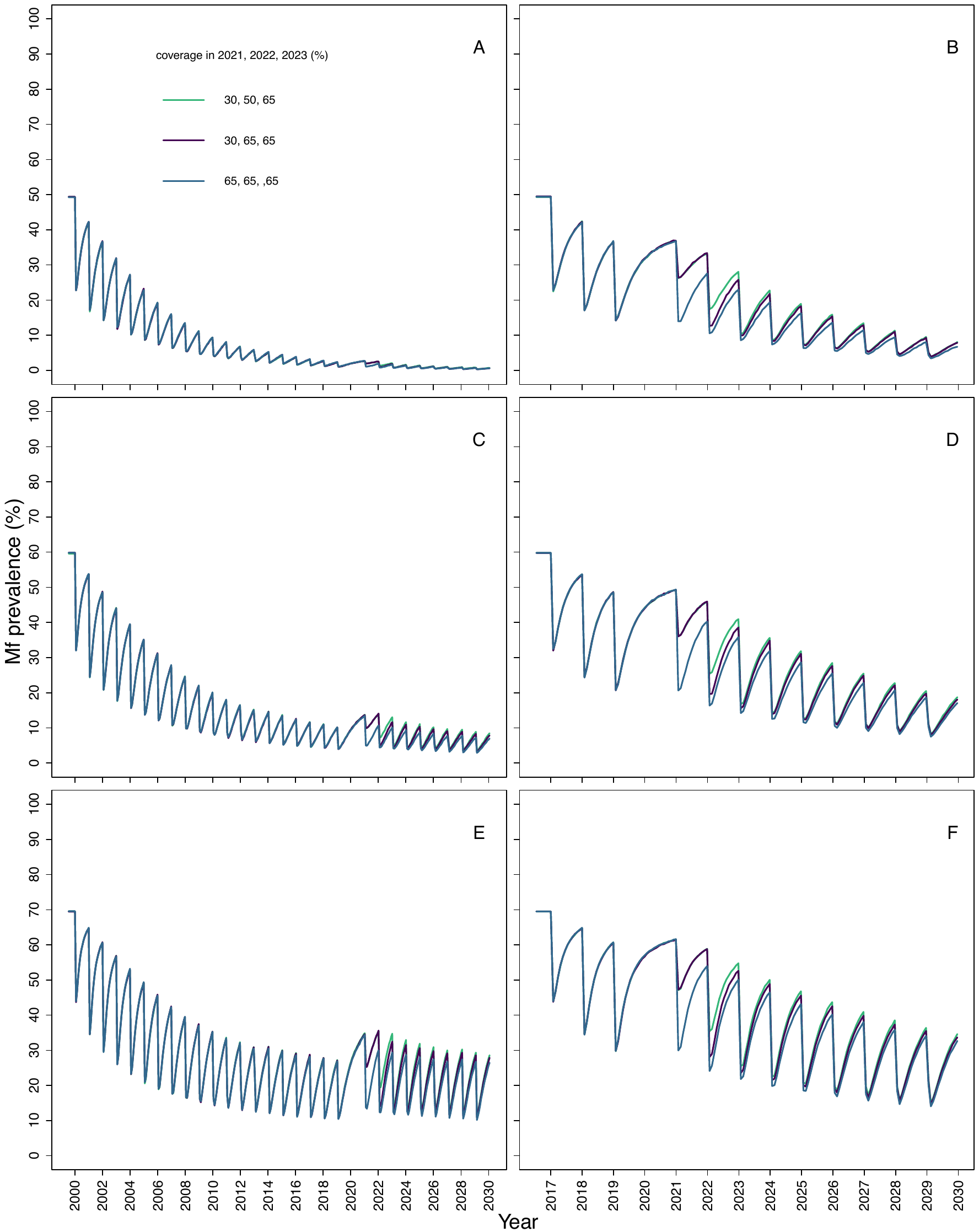

**Figure S2.** **Temporal dynamics of *Onchocerca volvulus* microfilarial (Mf) prevalence predicted by EPIONCHO-IBM assuming a 1-year interruption (2020) to annual mass drug administration (MDA) with ivermectin due to COVID-19 and no mitigation strategies, for three levels of coverage during treatment resumption.** The pre-intervention (baseline) microfilarial prevalence in individuals aged ≥5 years is 50% (**A, B**), 60% (**C, D**) and 70% (**E, F**). Annual MDA takes place from 2000 to 2030 (early-start programmes, **A, C, E**) or from 2017 to 2030 (late-start programmes, **B, D, F**), assuming no treatment in 2020 and no remedial strategies subsequently. The proportion of systematic non-participation is set to 5%.

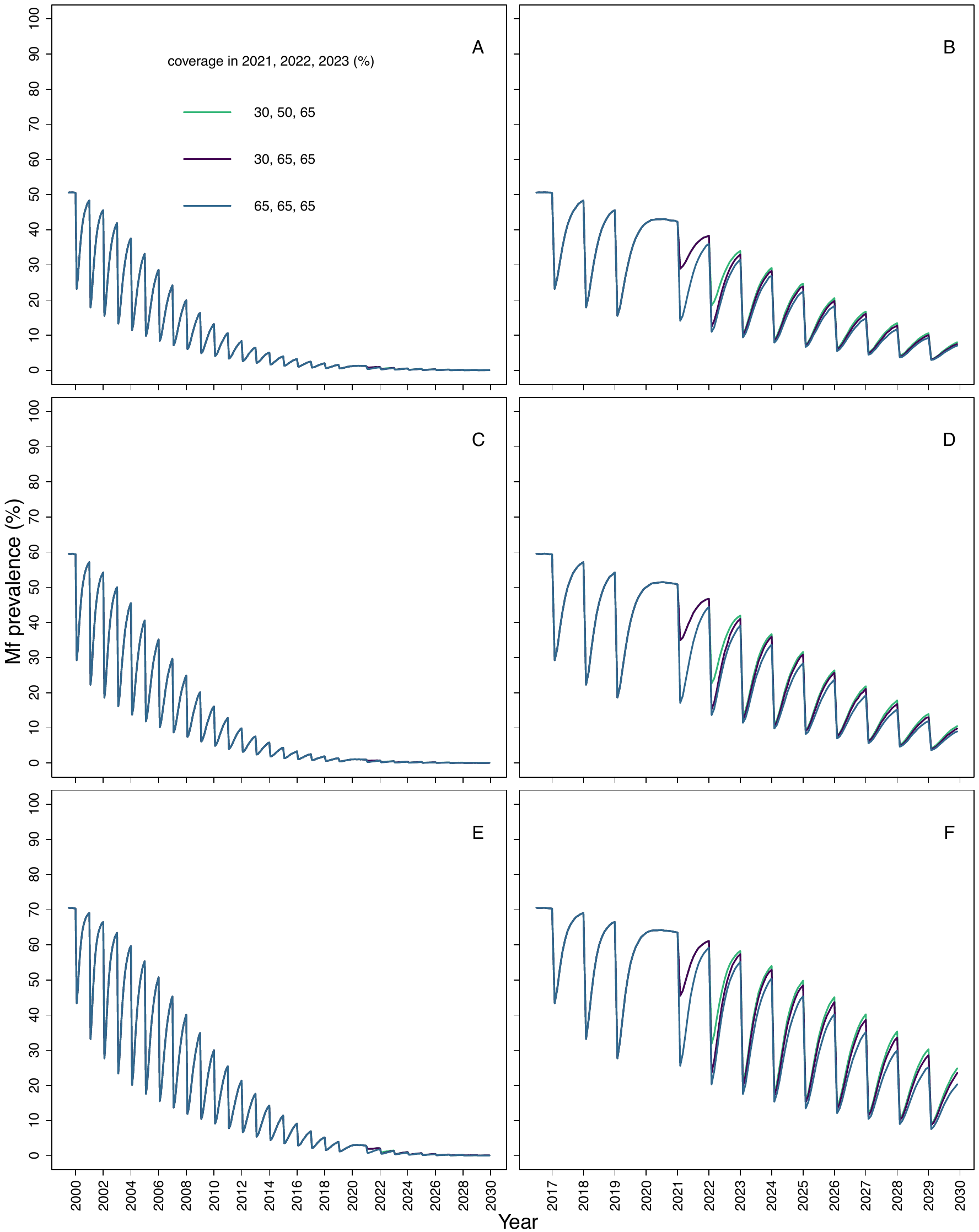
**Figure S3.** **Temporal dynamics of *Onchocerca volvulus* microfilarial (Mf) prevalence predicted by ONCHOSIM assuming a 1-year interruption (2020) to annual mass drug administration (MDA) with ivermectin due to COVID-19 and no mitigation strategies, for three levels of coverage during treatment resumption.** The pre-intervention (baseline) microfilarial prevalence in individuals aged ≥5 years is 50% (**A, B**), 60% (**C, D**) and 70% (**E, F**). Annual MDA takes place from 2000 to 2030 (early-start programmes, **A, C, E**) or from 2017 to 2030 (late-start programmes, **B, D, F**), assuming no treatment in 2020 no remedial strategies subsequently. The proportion of systematic non-adherers is set to 5%.

***Dynamics of microfilarial prevalence with and without treatment disruptions***

An illustration of how the dynamics following an interruption differ from those from without an interruption in EPIONCHO-IBM and ONCHOSIM is presented in Fig. S4.

The (absolute) differences in microfilarial prevalence in 2022, between the situation when there is no interruption and when a 2-year interruption is simulated, are more pronounced for programmes with a shorter treatment history (early-start) than for those with a longer one (late-start) for low and moderate baseline endemicities in EPIONCHO-IBM. However, for a given treatment history this difference in microfilarial prevalence in 2022 (between an interruption and no interruption) did not necessarily increase with the baseline transmission setting. The absolute difference in microfilarial prevalence between no interruption and a 2-year (2020–2021) interruption (measured in 2022) was found to change non-monotonically with baseline microfilarial prevalence, i.e., increasing up to intermediate baseline microfilarial prevalence and then declining in both models. Furthermore, the relative differences decreased with increasing baseline endemicity, although this was more pronounced for EPIONCHO-IBM than ONCHOSIM (Fig. S4).

Density-dependent constraints operating on parasite establishment within humans (assumed in EPIONCHO-IBM but not in ONCHOSIM) have non-trivial implications for observed patterns of bounce-back between treatment rounds, and also how the prevalence following an interruption to MDA differs from temporal infection dynamics in which there is no interruption. From one year to the next during annual MDA, the release of density-dependent constraints (following MDA) contributes to the more pronounced increases in microfilarial prevalence between rounds, predicted by EPIONCHO-IBM, than those predicted by ONCHOSIM. When comparing the microfilarial prevalence in 2021 and 2022 after an interruption (when transmission begins to increase), with the microfilarial prevalence in the same years without an interruption, the difference can *decrease* with increasing baseline endemicity due to the functional form of density dependence that has been assumed. The functional form of the density-dependent process assumed in EPIONCHO-IBM for parasite establishment within humans is roughly sigmoidal when plotted on a log scale, meaning that parasite establishment can decrease rapidly with increasing transmission or remain roughly constant.^1^ For lower transmission settings, which occupy flatter parts of this relationship, increasing transmission during an interruption is not associated with large decreases in parasite establishment. For higher transmission settings, which occupy decreasing (as transmission increases) parts of this relationship, increasing transmission during an interruption results in reduced parasite establishment. Since ONCHOSIM incorporates less density-dependent constraints than EPIONCHO-IBM (both models assume that they operate in parasite establishment within the vectors, but only in EPIONCHO-IBM these processes also operate in humans),^3^ these patterns are less pronounced in ONCHOSIM but still visible (Fig. S4).

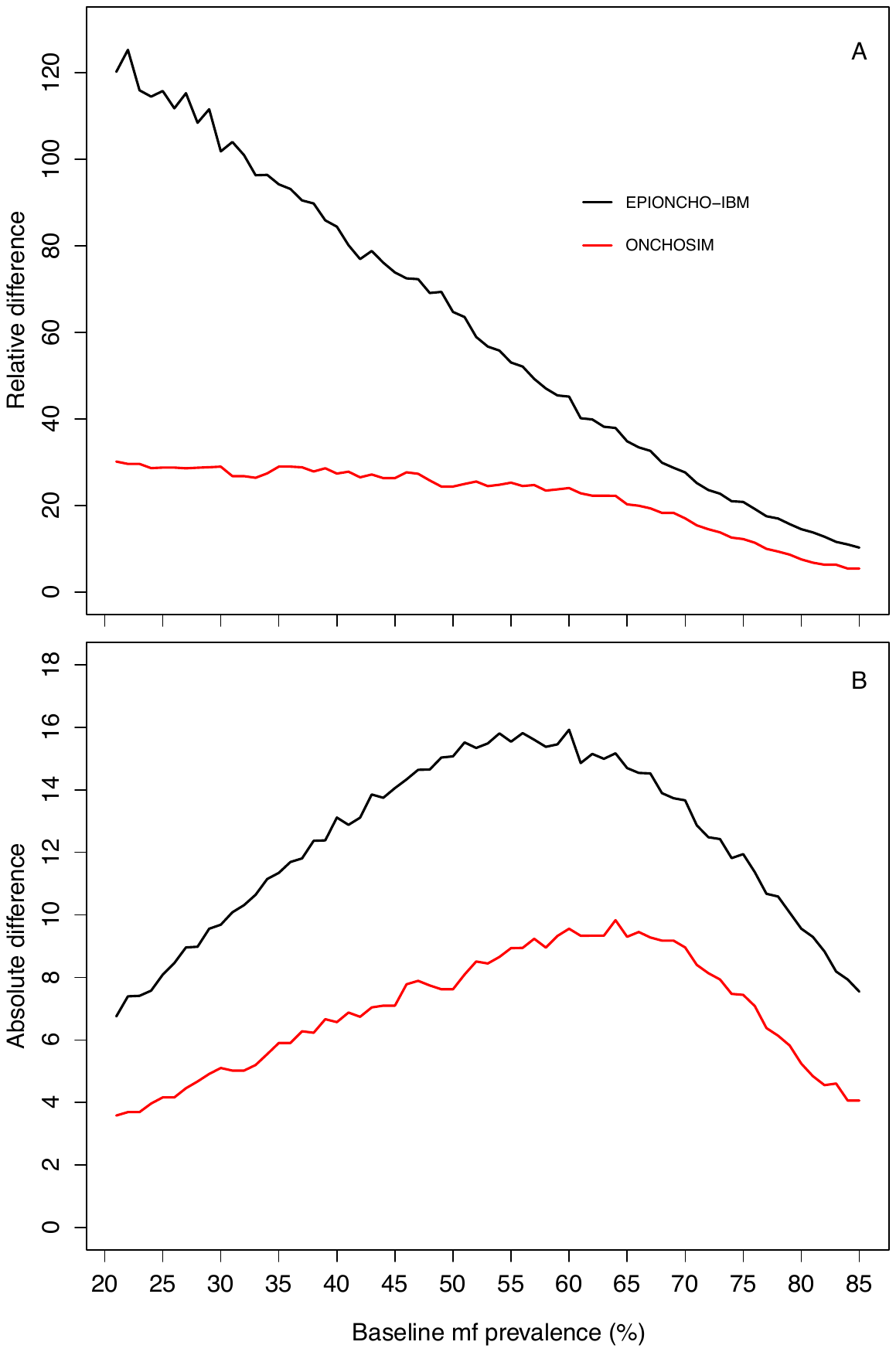

**Figure S4. The relative (A) and absolute (B) differences in microfilarial prevalence in 2022 between a 2-year interruption in MDA (2020 and 2021) and no interruption in MDA for EPIONCHO-IBM (black lines) and ONCHOSIM (red lines).** Annual ivermectin MDA is assumed to begin in 2017. The therapeutic coverage is assumed to be 65%, with the exception of 30% in 2022 following a 2-year interruption. The proportion of systematic non-participation is set to 5%.

***Influence of start year and baseline microfilarial prevalence on elimination probabilities***

The impact of a 1- and 2-year interruption on elimination probabilities with start years ranging from 2000 to 2020, and baseline microfilarial prevalence ranging from 20% (hypoendemicity) to 85% (holoendemicity) is presented, respectively, in Fig. S5 and S6.

Although in the African Programme for Onchocerciasis Control (APOC), MDA treatment was prioritised for areas with microfilarial prevalence ≥40% (i.e. meso- and hyperendemic areas), the range of baseline microfilarial prevalence in individuals aged ≥5 years explored in the simulations was set to 20% – 85% for the sake of completeness. In some former Onchocerciasis Control Programme in West Africa (OCP) countries, hypoendemic foci have also been treated, particularly when programmes have been integrated into wider-umbrella NTD remits that include ivermectin MDA for lymphatic filariasis.^9^

In Fig. S5, no interruption to MDA is represented by solid lines. A 1-year interruption (in 2020) is represented by dashed lines, assuming that coverage (of total population) from 2021 onwards i- 65%. The dotted lines assume that coverage in 2022 is 30%, increasing to 65% from 2023 onwards. In Fig. S6, no interruption to MDA is represented by solid lines and a 2-year (2020 and 2021) interruption by dashed lines. Because a 2-year interruption would represent a greater disruption to MDA programmes due to a prolongation of the COVID-19 pandemic, the therapeutic coverage in 2022 was assumed to be 30%. The proportion of systematic non-participation is set to 5% throughout all simulations.

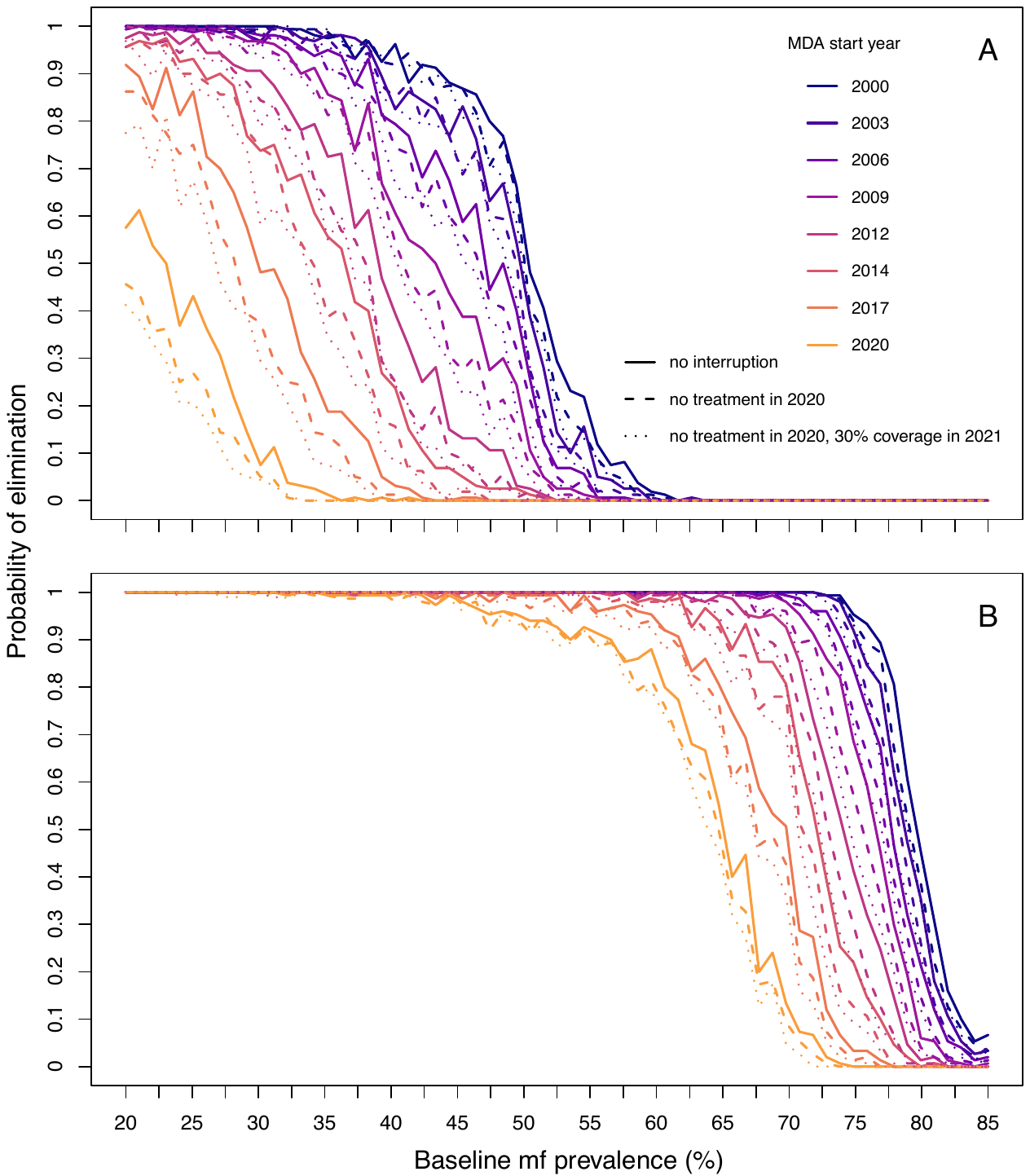

**Figure S5.** **Elimination probabilities versus pre-intervention (baseline) endemicity predicted by (A) EPIONCHO-IBM and (B) ONCHOSIM for a range of mass drug administration (MDA) histories with ivermectin with and without a 1-year (2020) interruption due to COVID-19 and no mitigation strategies.** Annual MDA programmes start in 2000, 2003, 2006, 2009, 2012, 2014, 2017, 2020 and finish in 2030. The range of baseline microfilarial prevalence in individuals aged ≥5 years is 20% – 85%. No interruption to MDA is represented by solid lines; a 1-year interruption is represented by dashed (65% coverage in 2021) and dotted (30% coverage in 2021) lines. The proportion of systematic non-participation is set to 5%.

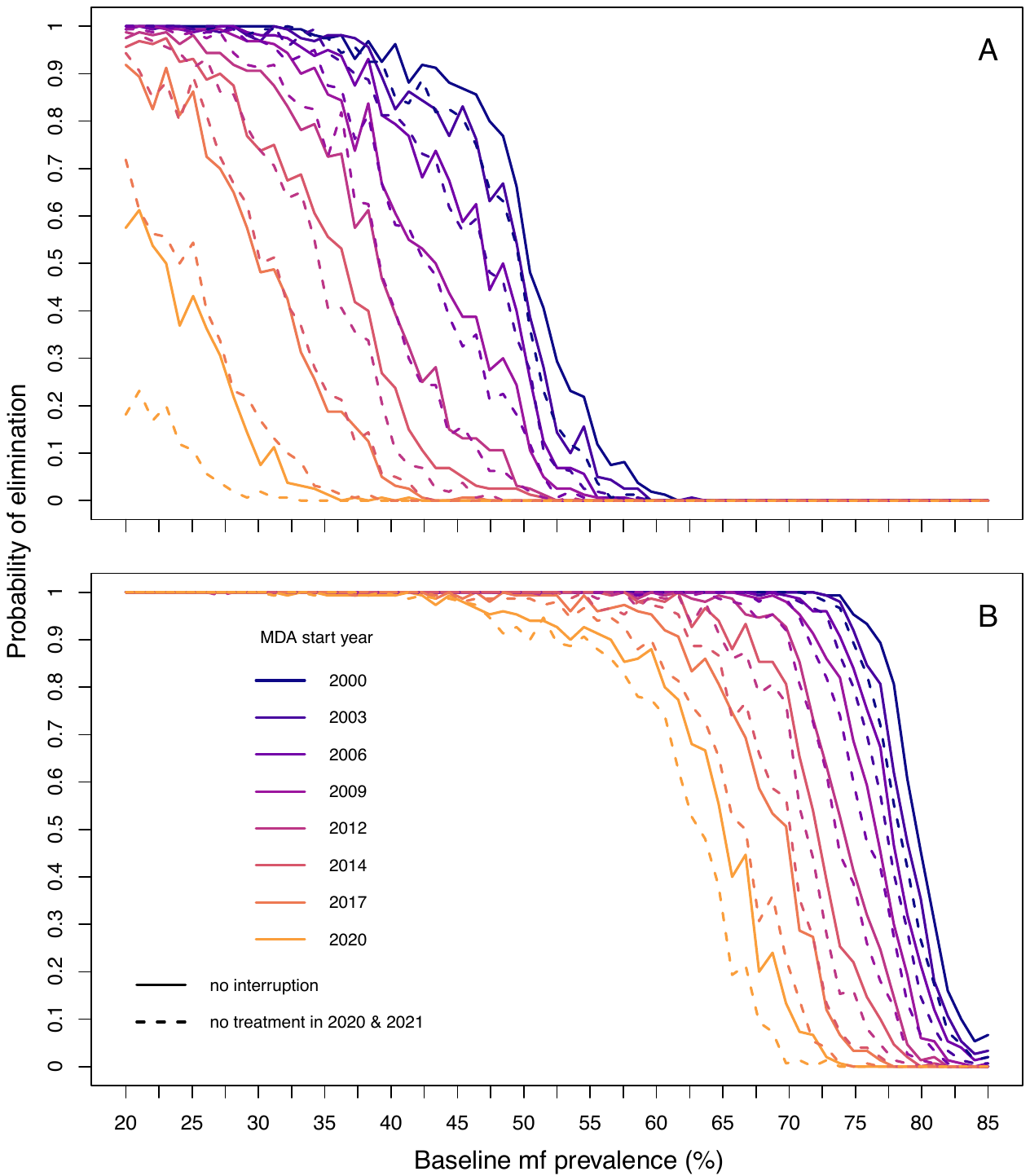

**Figure S6.** **Elimination probabilities versus pre-intervention (baseline) endemicity predicted by (A) EPIONCHO-IBM and (B) ONCHOSIM for a range of mass drug administration (MDA) histories with ivermectin with and without a 2-year (2020 and 2021) interruption due to COVID-19 and no mitigation strategies.** Annual MDA programmes start in 2000, 2003, 2006, 2009, 2021, 2014, 2017, 2020 and finish in 2030. The range of baseline microfilarial prevalence in individuals aged ≥5 years is 20% – 85%. No interruption to MDA is represented by solid lines; a 2-year interruption is represented by dashed lines. The therapeutic coverage is assumed to be 65%, with the exception of 30% in 2022. The proportion of systematic non-adherers is set to 5%.

***The impact of mitigation strategies***

**Microfilarial prevalence dynamics:** Two possible mitigation strategies were investigated to understand their influence on temporal dynamics of microfilarial prevalence. These strategies comprise either remedial biannual ivermectin MDA (i.e. increasing treatment frequency) or remedial coverage (i.e., increasing the therapeutic coverage of annual MDA). As these remedial strategies are more important following a 2-year (2020 and 2021) interruption, the results were presented in the Main Text. Here, we report their impact when the disruption to MDA only applies to 2020. The temporal microfilarial prevalence dynamics for a 1-year interruption with either remedial increased treatment frequency or therapeutic coverage are shown, respectively, in Fig. S7 and S8.

**Elimination probabilities:** For a 2-year interruption, although neither remedial strategy is sufficient to achieve the same elimination probabilities found in the absence of an interruption, 2 years of remedial biannual MDA results in higher elimination probabilities than remedial increase in coverage (Fig. S9). For some treatment histories and pre-intervention endemicities (which do not result in 100% elimination probability without interruption), 1 year of missed MDA followed by 1 year of remedial biannual MDA gives similar elimination probabilities to those in the absence of MDA disruptions (Fig. S10).

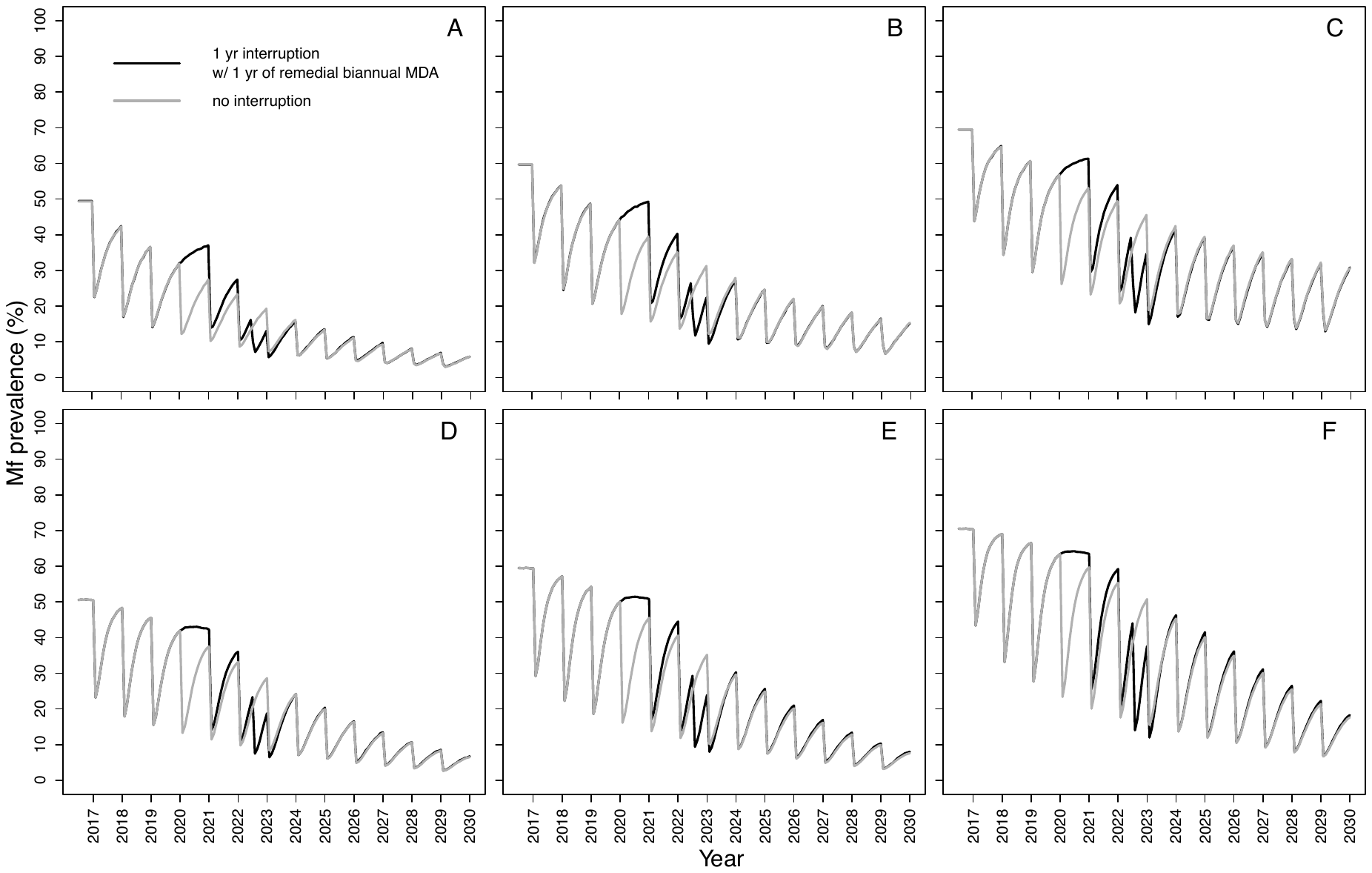
**Figure S7.** **Temporal dynamics of *Onchocerca volvulus* microfilarial (Mf) prevalence predicted by** **EPIONCHO-IBM (A – C) and ONCHOSIM (D – F) with a 1-year (2020) interruption, due to COVID-19, to mass drug administration (MDA) with ivermectin (2017 – 2030) and remedial biannual MDA.** The pre-intervention (baseline) microfilarial prevalence (in individuals aged ≥5 years) is: 50% (**A, D**), 60% (**B, E**), and 70% (**C, F**). Grey lines represent no interruption; black lines indicate no treatment in 2020 but with one year of remedial biannual MDA in 2022. From 2023 onwards, the frequency of treatment reverts to annual ivermectin MDA. The therapeutic coverage is assumed to be 65%. The proportion of systematic non-participation is set to 5% throughout all simulations.

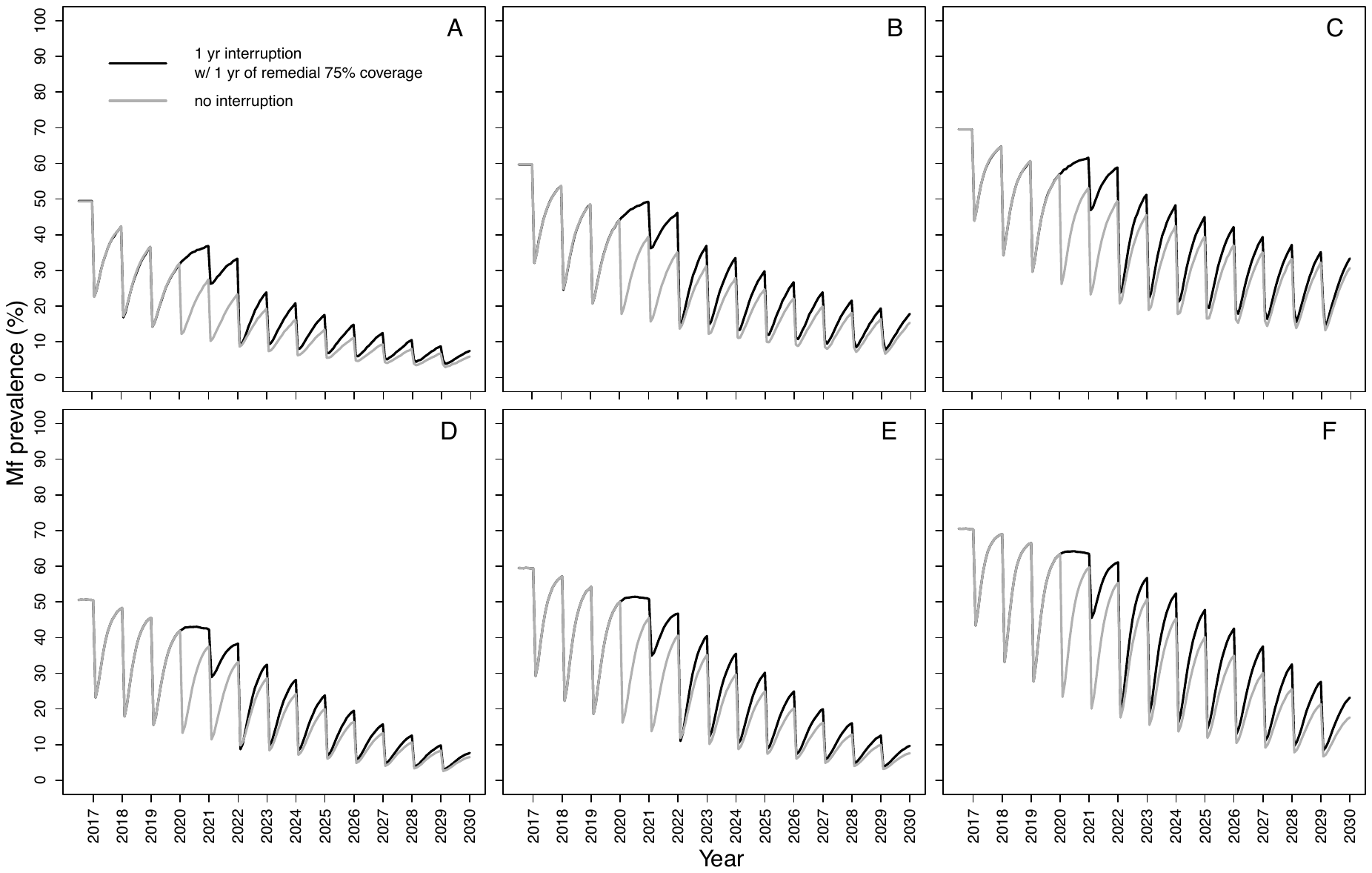

**Figure S8.** **Temporal dynamics of *Onchocerca volvulus* microfilarial (Mf) prevalence predicted by** **EPIONCHO-IBM (A–C) and ONCHOSIM (D–F) with a 1-year (2020) interruption, due to COVID-19, to annual mass drug administration (MDA) with ivermectin (2017–2030) and remedial annual high-coverage MDA.** The pre-intervention (baseline) microfilarial prevalence (in individuals aged ≥5 years) is: 50% (**A, D**), 60% (**B, E**), and 70% (**C, F**). Grey lines represent no interruption; black lines indicate no treatment in 2020 but with one year of remedial high-coverage MDA in 2022. The therapeutic coverage is assumed to be 65%, with the exception of 30% in 2021 (note this differs to our assumption for simulations with remedial biannual treatment) and 75% in 2022. From 2023 onwards, the therapeutic coverage of total population reverts to 65%. The proportion of systematic non-participation is set to 5% throughout all simulations.

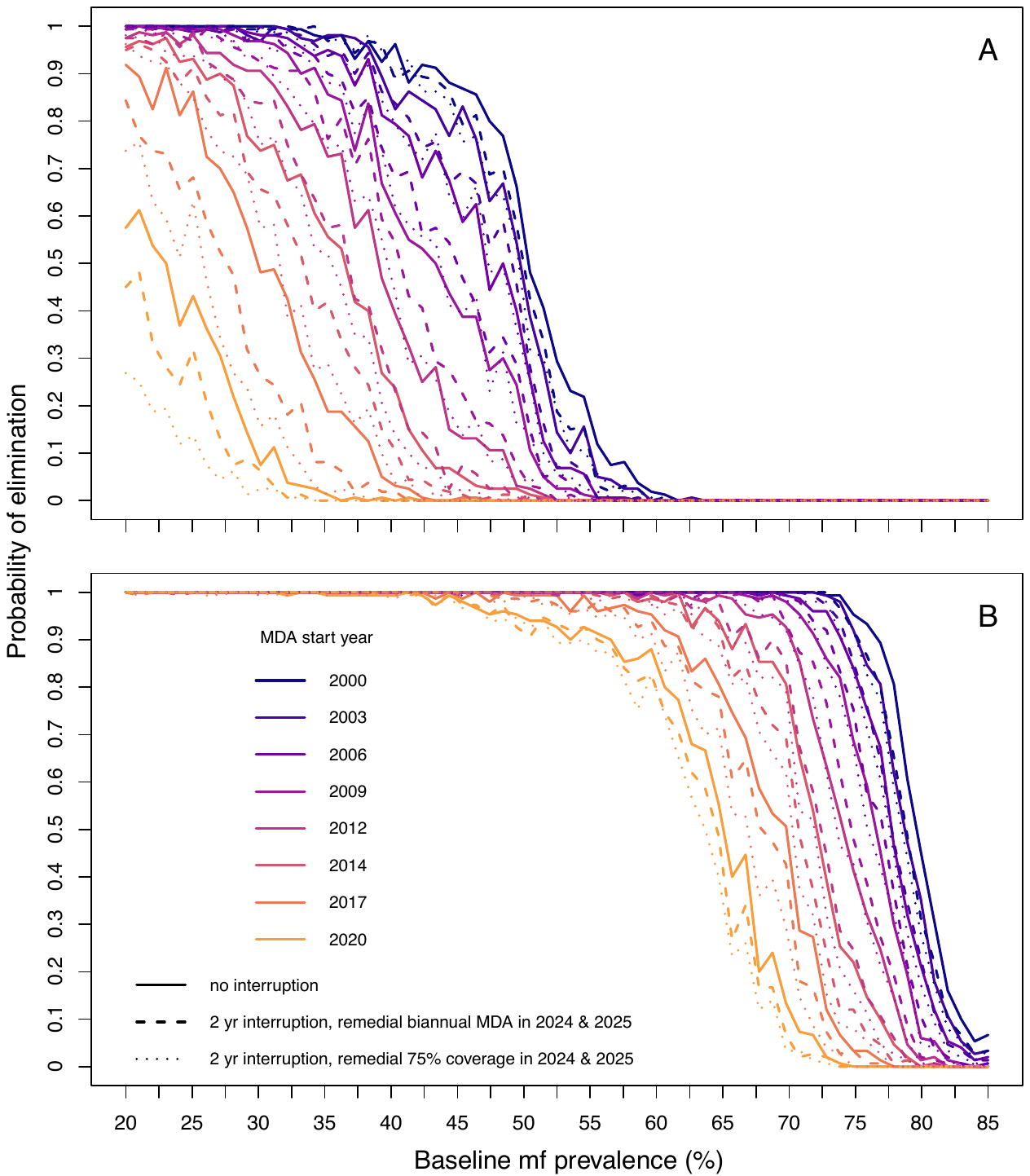

**Figure S9. Elimination probabilities versus pre-intervention (baseline) endemicity predicted by (A) EPIONCHO-IBM and (B) ONCHOSIM with a 2-year interruption to mass drug administration (MDA) with ivermectin (2020 and 2021) due to COVID-19 and two years of remedial biannual MDA or high-coverage annual MDA**. Annual ivermectin MDA programmes start in 2000, 2003, 2006, 2009, 2012, 2014, 2017, 2020 and finish in 2030; the range of baseline microfilarial prevalence (in individuals aged ≥5 years) is 20% – 85%. No interruption to ivermectin MDA is represented by solid lines; a 2-year interruption with increased treatment frequency (biannual MDA in 2024 and 2025) is represented by dashed lines; increased coverage of annual MDA (to 75% in 2024 and 2025) is represented by dotted lines. The therapeutic coverage is assumed to be 65% (of total population), with the exception of 30% in 2022. The proportion of systematic non-participation is set to 5%.

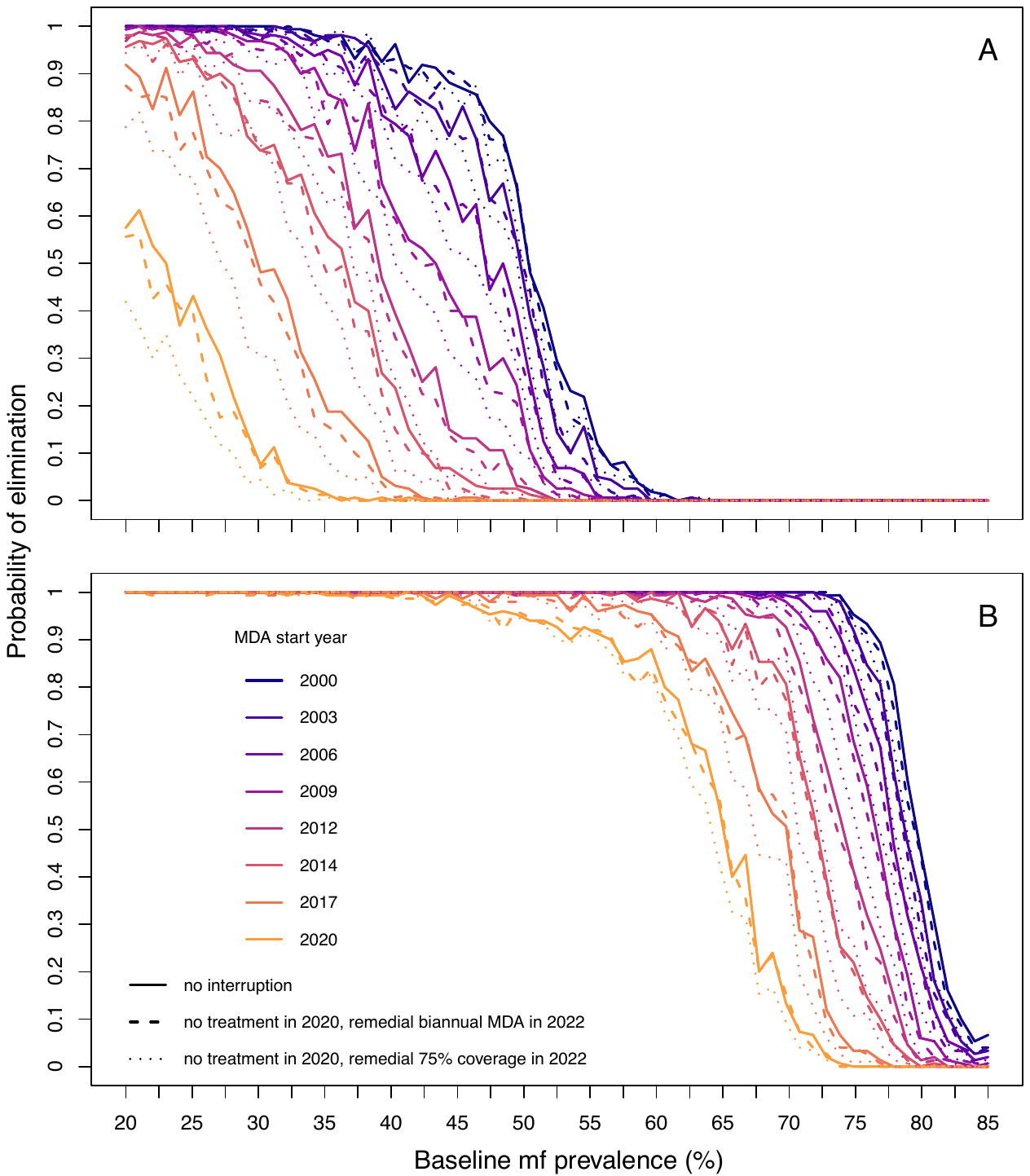

**Figure S10**. **Elimination probabilities versus pre-intervention (baseline) endemicity predicted by (A) EPIONCHO-IBM and (B) ONCHOSIM with a 1-year interruption to mass drug administration (MDA) with ivermectin (2020) due to COVID-19 and one year of remedial biannual MDA or high-coverage annual MDA**. Annual ivermectin MDA starts in 2000, 2003, 2006, 2009, 2012, 2014, 2017, 2020 and finish in 2030; the range of baseline microfilarial prevalence (in individuals aged ≥5 years) is 20% – 85%. No interruption to ivermectin MDA is represented by solid lines; a 1-year interruption with increased treatment frequency (biannual MDA in 2022) is represented by dashed lines; increased coverage of annual MDA (to 75% in 2022) is represented by dotted lines. The therapeutic coverage is assumed to be 65%, with the exception of 30% in 2021 when assuming remedial 75% coverage in 2022. We assume 65% coverage in 2021 before the implementation of remedial biannual MDA. The proportion of systematic non-participation is set to 5%.

***Individual-level microfilarial loads***

The insets provided in Fig. 7 of the main text illustrate the mean microfilarial load by age and sex. To provide an understanding of the individual microfilarial loads in addition to a visualisation of the mean, Fig. S11 plots, for EPIONCHO-IBM, the number of microfilariae per mg of skin for simulated individuals.

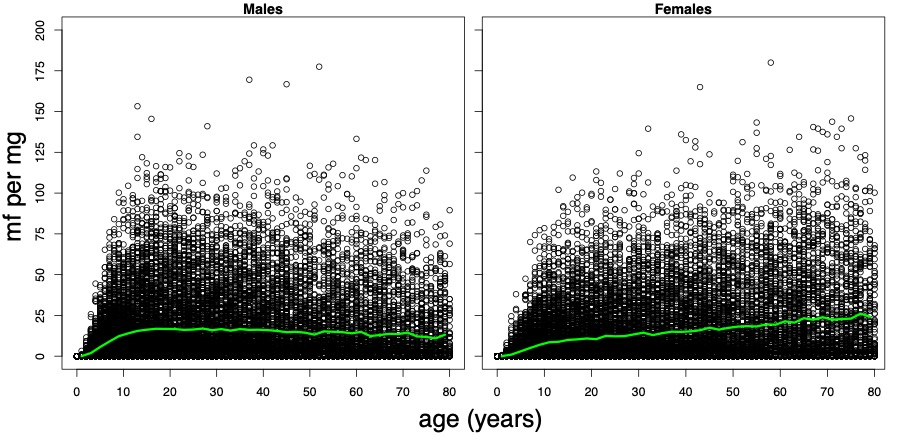

**Figure S11.** **Age- and sex-profiles of *Onchocerca volvulus* microfilarial intensity (number of mf per milligram of skin) predicted by** **EPIONCHO-IBM for males (left panel) and females (right panel) at baseline with a 70% microfilarial prevalence in individuals aged ≥5 years**. The black points represent individuals and the green lines represent the mean infection intensity within 1-year age classes. Simulated individuals are from 100 repeat simulations with population sizes of 440.
